# Mapping the electrophysiological structure of dystonic Globus Pallidus pars interna through intraoperative microelectrode recordings

**DOI:** 10.1101/2022.11.08.22281989

**Authors:** Ahmet Kaymak, Matteo Vissani, Sara Rinaldo, Roberto Eleopra, Luigi Romito, Alberto Mazzoni

## Abstract

**Objective:** The Globus Pallidus pars interna (GPi) is one of the main targets for Deep Brain Stimulation (DBS) therapies for dystonia and other movement disorders. Still, a complete picture of the spiking dynamics of the nucleus is far from being achieved. Microelectrode recordings (MER) provide a unique brain window opportunity to shed light on GPi organization, which might support intraoperative DBS target localization, as previously done for the Subthalamic nucleus (STN).

**Approach:** Here we propose a novel procedure to analyze explorative MERs from DBS implants in dystonic patients. The procedure identifies the neural activity markers discriminating neurons in the GPi from those in the neighbouring structures, as well as the markers discriminating neurons located in different regions within the GPi.

**Main results:** The identification of the borders of the GPi based on neural markers was a difficult task, due to internal inhomogeneities in GPi firing dynamics. However, the procedure was able to exploit these inhomogeneities to characterize the internal electrophysiological structure of the GPi. In particular, we found a reliable dorsolateral gradient in firing activity and regularity.

**Significance:** Overall, we characterized the spatial distribution of neural activity markers in the dystonic GPi, paving the way for the use of these markers for DBS target localization. The procedure we developed to achieve this result could be easily extended to MER performed for other disorders and in other areas.

## Introduction

Deep brain stimulation (DBS) of the internal segment of the globus pallidus (GPi) has been established as a highly therapeutic surgical procedure for severe idiopathic dystonia [1–3]. The inappropriate location of the active contacts explains most of the therapeutical failures in GPi DBS for dystonia, which increases the rate of revisions and remissions of previously implanted DBS leads [4,5]. Maximizing the overlap between the GPi optimal location and the 3D spread of the current delivered by the active contact of the DBS lead is critical to achieving clinically significant outcomes [6,7].

Although advances in imaging protocols have significantly improved preoperative GPi targeting, methodological limitations – such as low spatial resolution and possible brain shifts during the surgery – owe the utilization of up-to-five microelectrode recordings (MER) to intraoperatively validate the target location [8–10]. However, MERs can be time-consuming, which limits their use if an expert electrophysiologist is not available in the operating room. Scaling up the number of microelectrodes used during electrophysiological mapping allows for gaining more information, but also increases the risk of complications [11].

Multiple human intraoperative centres showed that MERs yield accurate electrophysiological spatial demarcation of target borders and identification of neural patterns that highlight optimal stimulation settings as well as discriminate clinical states [12–20]. Most of these studies focused on the Subthalamic nucleus (STN) *per se*, which represents another important DBS target for dystonia and other movement disorders, such as Parkinson’s Disease (PD) [21,22].

The peculiar characteristics of the transition from the external segment of the globus pallidus (GPe) to GPi, such as the sparseness of the neurons along the pallidum, the absence of a marked reduction in neuronal activity, and the high discharge rate variability, challenged the electrophysiological determination of GPi borders [27,28]. Occasionally, the identification of increasing background activity and *border cells*, characterized by low firing rates (2 – 20 Hz), may help identify the GPe-GPi transition.

Multiple MERs studies compared the activity of GPi neurons either across different diseases or before, during, and after functional microstimulation [24–31], neglecting or restricting the topographical analysis along the pre-planned trajectory axis. These studies identified *high-frequency cells* whose discharge rate (up to 150 Hz) may feature movement disorders. It is assumed that PD GPi cells are characterized by a higher discharge rate than dystonia GPi cells. Low-frequency oscillatory activity (2 – 10 Hz) and bursting patterns are also observed in GPi across different diseases. All these findings may be confounded by different anaesthetic protocols and the heterogeneity of neural features (both discharge rate and discharge pattern) in different Dystonic phenotypes. Strikingly, a topographic map of the spiking dynamics of GPi neurons has not been elucidated yet. Clinically-informed decisions may leverage these topographic maps to define relationships between neural activity and optimal stimulation sites.

Interestingly, complementary approaches using postoperative local field potential (LFP) recordings showed excessive entrainment of spatially localized theta oscillations in the Dystonic GPi, indexing the optimal stimulation site [32].

Here, our aim is to provide an MER-based machine-learning pipeline that retrospectively demarcates electrophysiologically-defined GPi borders and builds a topographic map of the internal GPi spiking dynamics in Dystonic patients. This work is in line with the growing interest in electrophysiological GPi physiomarkers and might lay the bedrock of the implementation of future computer-aid systems that support and hasten clinical decisions during the placement of the DBS lead in GPi DBS dystonic patients [17,32,33]. The development of algorithms that interpret short segments of MERs during DBS surgery is crucial to maximally exploit the limited time in operating room settings.

## Methods

### 2.1 Subjects

The research activity was conducted with seven male and three female patients who are diagnosed with primary dystonia. All the patients who took part in this study had severe dystonia, which significantly limited their ability to carry out their daily tasks. Patients were addressed to surgery relying on these inclusion criteria: diagnosis of primary generalized or multisegmental dystonia, normal behavioural, psychiatric, and cognitive profile, normal brain MRI and lack of response to medical treatment including anticholinergics, benzodiazepines, neuroleptics, baclofen and/or botulinum toxin injections. Patients underwent bilateral implantation of quadripolar electrodes, after giving their written informed consent to the procedure.

### 2.2 Surgery

The patient underwent frameless surgery. Preoperative images (MRI and CT with the fiducials on the skull) and planning were acquired the day before surgery. Non-stereotactic MRI brain scans (T1 with gadolinium and T2; slice thickness: 1.5 mm; without gap or overlap) were performed and, after the MRI acquisition, metal screw fiducials were fixed to the skull of each patient under local anaesthesia. Thereafter, a volumetric non-contrast scan (axial, slice thickness from 0.75 to 1 mm) with the fiducials in place was performed. For all the procedures, CT data were matched with the previous MRI datasets in the Medtronic Stealth Station (Framelink®; Medtronic, Minneapolis, MN).

To define the anatomic target, we used fixed distances from the midpoint of the AC-PC line on T1 MRI slices combined with the direct visualization of the nucleus on T2 MRI images, to identify the brain target (GPi). In our study, we did not have medical imaging that belongs to GPi DBS surgeries of patients to inspect the boundaries of GPi. Hence there is no certainty in terms of relative positions of MER trajectories to GPi for each subject. To overcome this issue, the DISTAL atlas[34] which is a three-dimensional subcortical brain atlas that also contains the anatomical position of GPi in Montreal Neurological Institute (MNI) space. Target coordinates derived from the stereotactic atlases registered to the mid-commissural point were manually entered into the planning program screen and identified onto the corresponding image slice. The atlas coordinates for the GPi nucleus were 21 mm lateral, 2 mm anterior, and 4 mm deep to the mid-AC-PC point. The anatomic coordinates of the preoperative target were finally defined by slightly adjusting the atlas coordinates and carefully evaluating details about the anatomic structures surrounding the estimated target, appreciable as a result of CT fusion with MRI. GPi definition was used later to determine whether sorted neurons are situated inside or outside of the GPi.

The patient was placed on the operating table, and the surgeon did the first navigation and marked the frontal entry points on the skin using a “passive frame”: referring to the fiducials screwed in the skull and assessing the correct position of the camera in the operating room, navigation on Stealth Station showed “geometry error”: geometry errors of the Nexprobe and the reference arc were less than 0.5mm in all the patients of this sample.

A drill hole of 14 mm in diameter was created 2.5 to 3.5 cm off the midline anterior to the coronal suture, and it was closed by a cap of fibrin sealant to avoid deliquoration. The NexFrame System® (Medtronic, Minneapolis, MN) was used for each procedure performed.

In the majority of the patients, surgery was performed while the patient was awake but, in those patients with severe dystonic fixed postures or those unable to tolerate an awake surgical procedure we performed surgery under general anaesthesia with a ketamine-based anaesthetic protocol. This anaesthetic protocol did not alter the electrophysiologic basal ganglia activity, allowing MER to be achievable and reliable during DBS surgery[35–37].

The choice of final electrode position was based on multi-electrode recording qualitative evaluation by an expert neurophysiologist, clinical efficacy during macro-stimulation, and the absence of significant macro-stimulation-induced collateral side effects. MER and macro-stimulation were performed using the Leadpoint System® (Medtronic, Minneapolis, MN). Finally, surgery was completed with the implantation of the subcutaneous extensions and the IPG in the sub-clavicle area.

### 2.4 Electrode Localization

The day after the surgical procedure, the patient underwent postoperative volumetric non-contrast scans (axial, slice thickness 0.75-1 mm) CT scans, to evaluate the final lead position and assess postoperative pneumoencephalus and rule out asymptomatic haemorrhage. The postoperative CT scan was merged with the preoperative CT and MRI images using Medtronic Stealth Station (Framelink®; Medtronic, Minneapolis, MN) to blend the AC-PC coordinates and angles of the trajectory of the final DBS leads. Each final lead location was identified as the centre of the beam-hardening artefact representing the deepest of the electrodes. This mapping of MER electrode trajectories to MNI space was a rigid transformation.

### 2.5 Electrophysiological recordings

Electrophysiological explorative microelectrode recordings were simultaneously recorded along five different trajectories: anterior, lateral, medial, central, and posterior trajectories. The recordings were collected bilaterally and simultaneously from the right and left hemispheres with varying depths from -10 mm to +3.5 mm (0 mm is the planned target point) by using FHC semi-micro-electrode (impedance=1 MOhm (FC2002, Medtronic R, Minneapolis, MN)). During the recording procedure, an analogue 500 Hz high pass filter was applied. MERs were sampled at 24000 Hz for 10 seconds. In both hemispheres, the MER trajectories are parallel to the GPi’s dorsoventral axis, with the central trajectory crossing the DBS target (see Supplementary Material Section 2.1).

We examined 1640 raw microelectrode recordings of 10 patients (min. 99, max. 246, 164±50.19 recordings per subject) (see Supplementary Material Section 2). Putative single unit activities (SUAs) are successfully isolated from raw MERs (Figure 1A). SUA represents the section of the raw data that belongs to the spiking of a single neuron. The [-0.5 +2.5] ms time interval around the peak values of all spikes extracted from raw data and stored as the neural activity of a single neuron[38].

**Figure 1.**
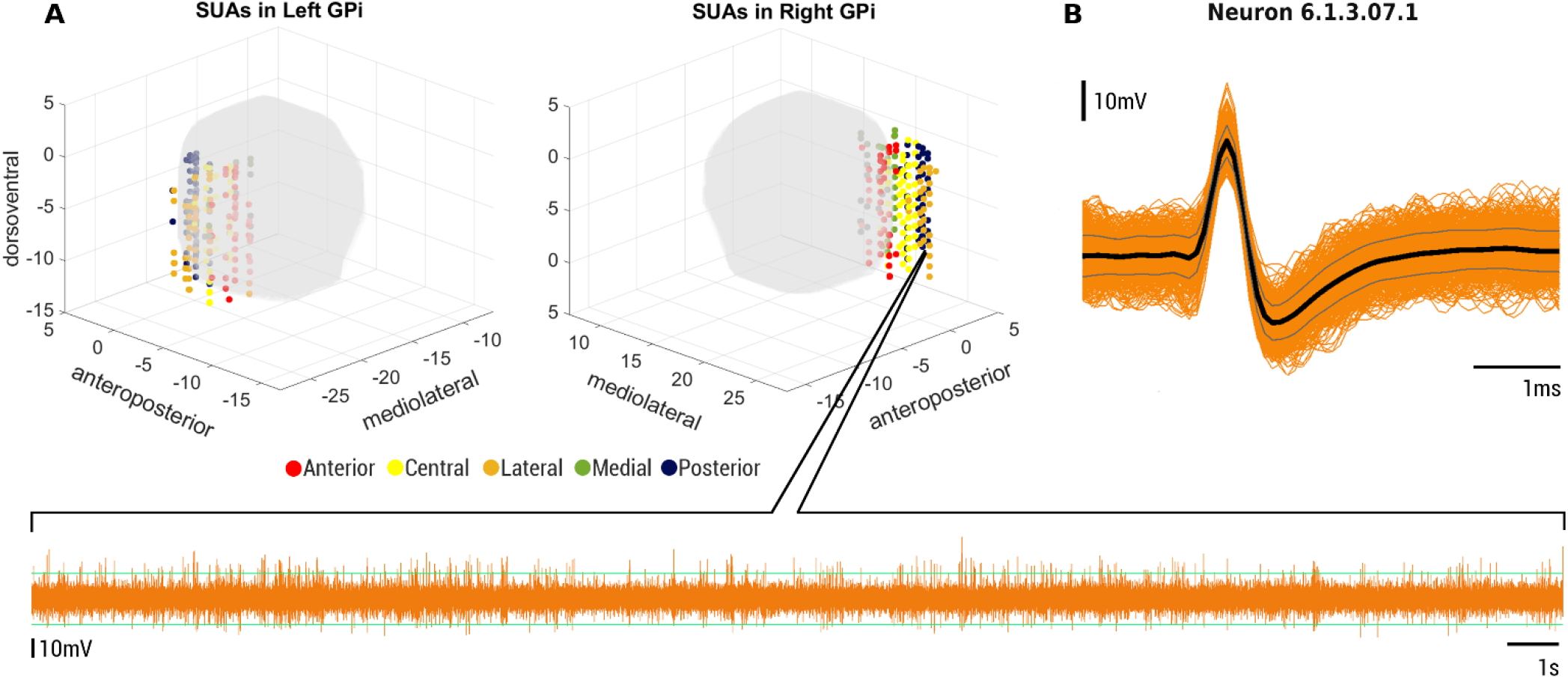
Isolated single unit representations on GPi in MNI reference frame. **(A)** Locations of the single-unit activities are displayed in two dystonic GPi based on their MNI coordinate definitions. Color-coded perpendicular lines along the dorsoventral axis of GPi represent electrode trajectories (anterior, central, lateral, medial, and posterior) GPi is depicted as defined by the DISTAL atlas. **(B)** In addition, a raw MER recording and its single-unit activity from -7mm (lateral trajectory) in the right hemisphere of patient 6 are presented.

### 2.6 Computational pipeline

To analyse MERs, we implemented a computational pipeline where each module is specialized for a particular task. In total, we implemented four modules: (1) Offline Spike Sorting Module, (2) Candidate Biomarker Generation Module, (3) Mutual Information Analysis Module, and (4) Machine Learning Module (see Figure 2).

**Figure 2.**
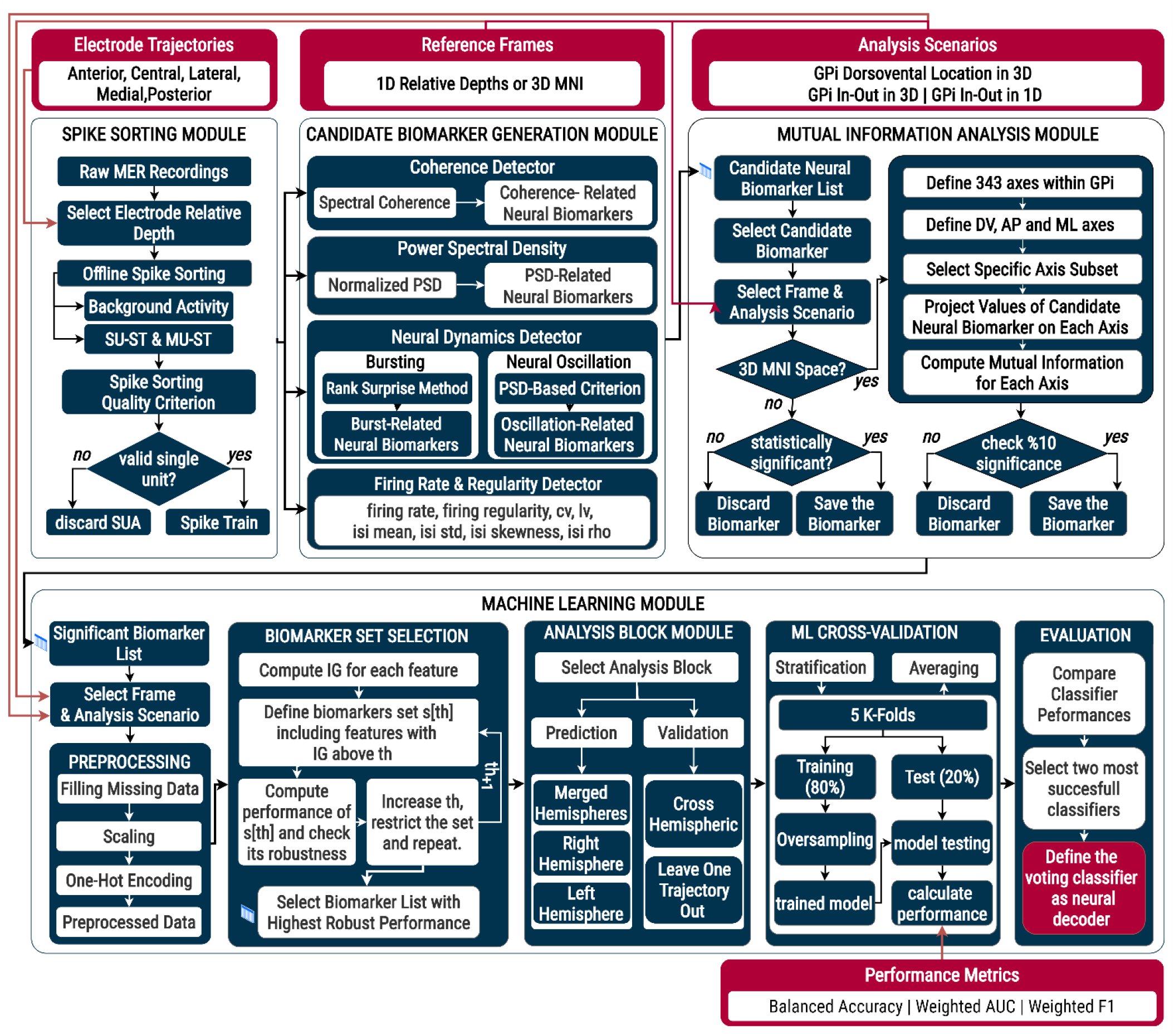
Modular structure of neural decoder pipeline for neuron localization. The computational pipeline of the single-unit activity localization is based on the use of candidate neural biomarker definitions in machine learning algorithms. The pipeline is made up of four different modules: Spike Sorting Module, Candidate Neural Biomarker Generation Module, Mutual Information Analysis Module, and Machine Learning Module. Abbreviations: AP, anteroposterior; DV, dorsoventral; IG, information gain; ML, mediolateral.

#### 2.6.1 Spikes detection and sorting

In the offline spike sorting module (Figure 2, top, left box), spike detection and sorting operations were conducted (see Supplementary Material Section 3.2). For spike identification and sorting, we used Wave Clus MATLAB ToolBox[39]. Double-thresholding (4 standard deviations from the baseline signal) was used to designate spike peaks, and 64-wavelet coefficients were computed for each potential spike. Then, using a superparamagnetic clustering method, the spikes are clustered to specific neurons based on their wavelet coefficients. We decomposed raw recordings into three main components: single-unit activity (SUA), multi-unit activity (MUA), and background activity (BUA)[38](see Supplementary Material Section 3.3)[14,15]. Each of these components carries different types of information about surrounding neural activity[38]. Noteworthy, we applied additional stability criteria that helped us prevent the inclusion of candidate neurons that do not reliably resemble SUA signatures for further analysis (see Supplementary Material Section 3.4).

#### 2.6.2 Reference frames

We carried out all of our analyses in two different reference frames: 1D relative depth and 3D MNI reference frames. The definition of two distinct reference frames allowed us to utilize our localization tasks both for clinical aspects and for understanding the electrophysiological structures of GPi. The 1D reference frame denotes the direction that is primarily parallel to GPi’s dorsoventral axis. During the GPi DBS surgeries, neurosurgeon teams penetrate the electrodes perpendicularly to GPi. Localization and real-time feedback in the 1D reference frame offer clinical utility from this standpoint.

Although the 1D reference frame has clinical relevancy, it falls short of accurately reflecting neural dynamics in terms of the actual anatomical structure of GPi. To address this problem, we used the MNI coordinates of SUAs to characterize the pathological dynamics of dystonic GPi and tried to localize neurons in the 3D space.

#### 2.6.3 Candidate neural biomarkers

Even if recent advances have been made in GPi bordering[40,41] and functional segregation of GPi [37], these approaches are not adequate to provide real-time feedback to neurosurgeon teams during DBS procedures for target localization. We used the definition of candidate neural biomarkers to locate and characterize neural behaviour inside different areas of GPi and to distinguish GPi borders for patients with primary dystonia. Operatively, we categorized neural dynamics into five different groups: (a) firing rate and regularity, (b) neural oscillations, (c) neural bursts, (d) spectral coherence between SUA and BUA, and the power spectrum of single-unit activity in different frequency bands. Then in each group, we defined multiple candidate neural biomarkers that represent a particular aspect of the related neural phenomena (see Supplementary Material Section 4).

#### 2.6.4 Power Spectrum Estimation

Firstly, we extracted the action potentials of single-unit activity from raw MER recording and remove the mean to get rid of the power at f=0 Hz. In the following step, we removed the power line noise (50 Hz) and its first five harmonics based on spectrum interpolation technique to prevent introducing bias to our power spectrum and neural oscillation estimation[42]. We adopted the Welch PSD estimation by selecting the Hanning window as our tapering function with a window size equal to the data length divided by 20. The overlapping of the windows is selected as 0.5. In the last step of power spectrum estimation, we normalize the power spectrum by dividing the power values in each frequency by the total power of the SUA. This normalization procedure is essential for acquiring a comparable power spectrum across all isolated single units.

#### 2.6.5 Information analysis

Mutual information is a metric that endeavours to quantify the non-linear relationship between two variables. Briefly, it measures the decrease in entropy of one random variable by observing another random variable[43]. In this module (Figure 2, top, right box), we want to measure the relationship between candidate neural biomarkers and their position along with the two aforementioned reference frames (Section 2.6.2. and Figure 3). This entails computing the mutual information I(B,S) between the value of the biomarker and the position of the recording (1).

**Figure 3.**
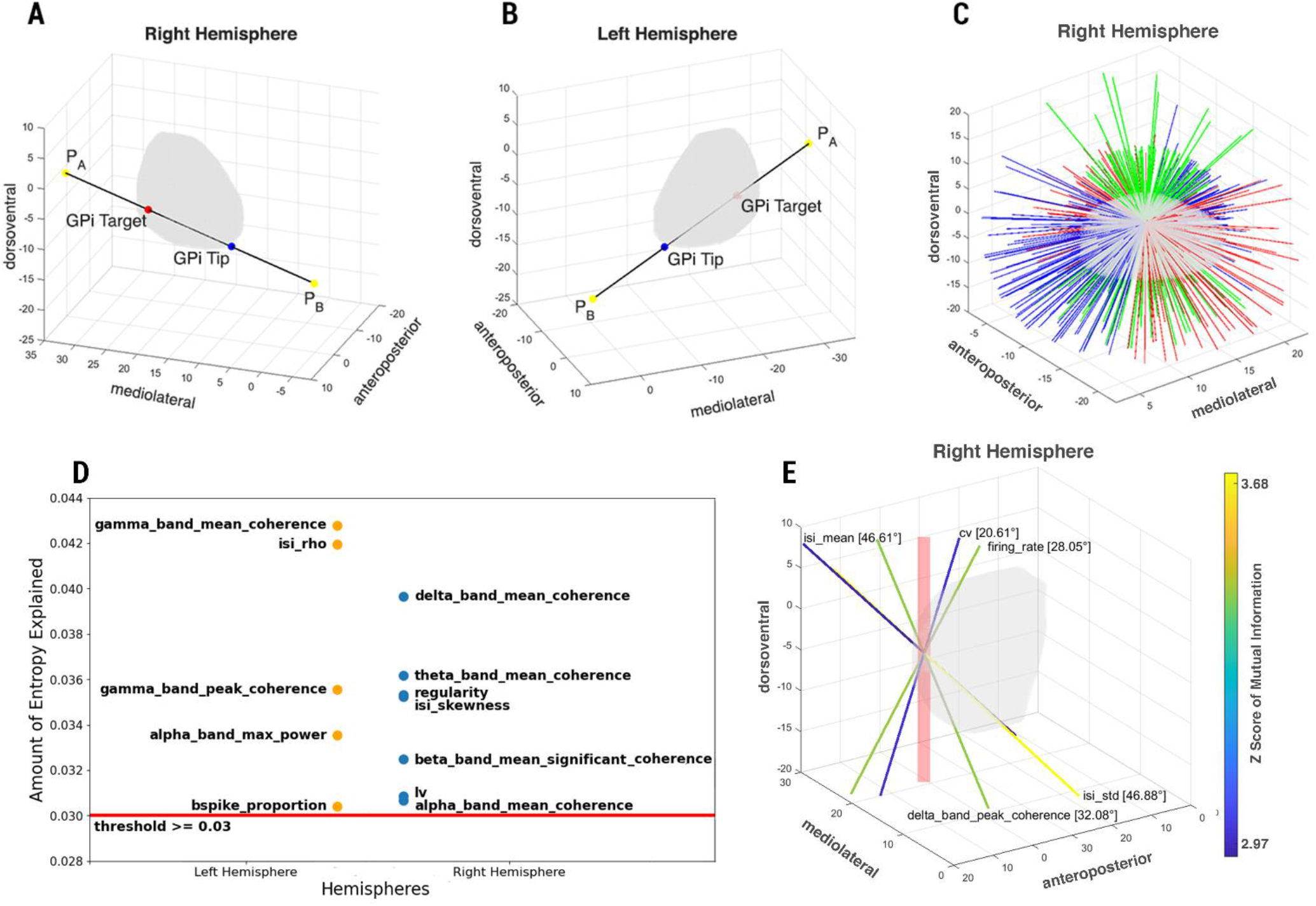
The initial direction definitions in both hemispheres and the lines scanning right hemisphere GPi for mutual information calculation in MNI space. **(A)** The initial line in the right hemisphere GPi that pass through *GPi*_*tip*_ and *GPi*_*target*_ and lies between *P*_*A*_ and *P*_*B*_ tips. **(B)** The initial line in the left hemisphere GPi that pass through *GPi*_*tip*_ and *GPi*_*target*_ and lies between *P*_*A*_ and *P*_*B*_ tips. **(C)** Directions that are generated based on the rotation of the initial line by Euler angles with 30-degrees steps in x, y and z axes to scan the whole GPi. The colour of directions represents the directions that belong to a specific anatomical axis. **(D)** The significant neural biomarkers for GPi in-out classification in 3D MNI reference frame for right and left hemisphere localization tasks in prediction block (mean explained entropy threshold ≥ 0.03). **(E)** The preferential directions of significant five neural biomarkers for dorsoventral localization in 3D MNI reference frame for merged hemispheres task of prediction block. The numeric values denote the angle between the preferential direction of the neural biomarkers and the dorsoventral anatomical axis of GPi. The red column represents the main direction of electrode trajectories that was used during GPi DBS surgeries.

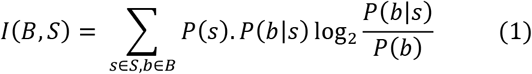

Where B is the set of values of the biomarker and S is the set of recording positions (in terms of relative depth level for the 1D reference frame and MNI coordinates in 3D MNI reference frame localization scenarios). P(b) is the probability over all recording sites of the biomarker having value b. The continuous values of the neural biomarker(b) were binned into four levels and represented with their corresponding bin values. P(s) is the probability of having a recording from position s over all biomarker values (which, by design, is the inverse of the number of recording positions). P(b|s) is the conditional probability of observing the biomarker value of a neuron at spatial component s.

Probability estimates during mutual information calculation can be impacted in the case of a limited amount of data. Since information is computed as the difference between two entropies, entropy bias is a significant issue here. The difference between the two entropy values is generally quite smaller in both entropies. Hence, even a tiny proportionate bias error in the entropy can have a significant impact on the information. As a result, a bias correction procedure should be applied to the calculation of mutual information. We chose the Panzeri–Treves[44] strategy out of all the bias correction approaches to tackle the sampling bias difficulties, and we computed the mutual information for each candidate neural biomarker. The significance of computed mutual information was assessed by generating a null distribution with the bootstrap test with 500 iterations. Based on the z-score of the computed mutual information, we defined the significance value (z≥2 for p≤0.05). We used the Information Breakdown Toolbox[45] in MATLAB to compute all the aforementioned information values.

For the 1D reference frame, mutual information is defined along electrode trajectories, and we only selected biomarkers that exhibit significant mutual information along these trajectories. Initially, we considered the depth level of neurons as the spatial location and the corresponding value of candidate neural biomarkers as the response for information analysis. Then we sorted and discretized the values of candidate biomarkers based on four equally populated bins. We had fifteen depth levels for the 1D reference frame.

In the 3D MNI reference frame, we determined the neuron locations based on matching the MER recording sites with their corresponding MNI coordinates (see Section 2.2.).

For utilizing the mutual information in 3D MNI space as a way of a feature reduction for our neural decoders, similar to the 1D reference frame, we adopted the approach of Mosher et al[19]. As opposed to their work, we did not employ principal component analysis (PCA) for feature reduction. We translated their optimal axis description to our preferential directions of candidate neural biomarkers definition. Instead of correlating the values of candidate biomarkers on rotated axes with anatomical positions of neurons, we used the mutual information to also grasp more complex non-linear relationships between the value distributions of candidate biomarkers and the MNI coordinates of neurons.

To define the preferential direction, i.e., the direction with the highest significant mutual information, of candidate neural biomarkers, we scanned the whole GPi. We defined two initial reference lines for the GPi scanning process. For the right hemisphere, we determined a line passing through the average target point of GPi DBS for the right hemisphere and the most lateral & anterior tip of GPi. Along this line, we define two tip points *P*_*A*_ and *P*_*B*_ that has a length three times the distance between *GPi*_*tip*_ and *GPi*_*target*_ points (Figure 3A). Similarly, for the left hemisphere, we defined another line passing through the target point of GPi DBS and the most medial & posterior tip of the left hemisphere GPi. Again, we defined two tip points *P*_*A*_ and *P*_*B*_ (Figure 3B). We rotated these initial lines with Euler angles from 0 to 180 degrees by 30-degree steps in the x, y, and z axes. Hence, this scanning procedure resulted in 343 rotated directions inside GPi (Figure 3C). On each of these 343 lines, we projected the neurons according to their MNI locations. Following the projection procedure, we split each line into 15 equally sized sections due to having a similar number of divisions with depth levels of electrode trajectories in the 1D reference frame. Therefore, the section on the selected direction that coincides with the projection of neuron location is used for mutual information calculation with the biomarker values of neurons in the 3D MNI reference frame.

To be more rigorous for the localization tasks that occur in a specific anatomical axis of GPi (i.e., dorsoventral localization of neurons within GPi), we did not consider mutual information results in all defined directions. Instead, we grouped these predefined 343 directions into three subsets. To achieve this division, we calculated the angle between each direction and all three anatomical axes that have the origin at the *GPi*_*target*_ point. Then we assigned each direction to the anatomical axis that has the smallest angle with it. With this approach, we defined subsets of directions that are aligned mostly with a specific anatomical axis of GPi. The red(N=112), green(N=111), and blue(N=120) coloured directions in Figure 3C indicate the subsets of direction that belong to the anteroposterior, dorsoventral, and mediolateral axes of GPi respectively. We utilized these three groups as follows: for the dorsoventral localization task, we only considered candidate neural biomarkers that show significant mutual information in at least 10% of dorsoventral directions. Thus, we evaluated the mutual information only in the relevant anatomical axis for each localization task and ignored the remaining directions.

We exploited the results of mutual information analysis as the first step of the feature selection mechanism for the machine learning module. The detailed information regarding the mutual information computation for different tasks can be found in the supplementary material.

#### 2.6.6 Machine learning decoding algorithm

The last module of the computational pipeline (Figure 2, bottom) is the machine learning module that contains the implementation of multiple neural decoders and cross-validation approaches to localize neurons in two different reference frames (1D and 3D). Apart from different reference frame definitions, we specified two analysis blocks: prediction and validation blocks. In terms of the prediction block, we conducted merged-hemispheres, right-hemisphere-only, and left-hemisphere-only classification tasks. To cross-validate the performances of trained models in the prediction block, we developed seven cross-validation classification tasks in the validation block. Firstly, we implemented five leave-one-trajectory-out validation tasks to check the performance of the merged-hemispheres model for each trajectory separately. We trained the merged-hemisphere model with neurons of four trajectories and completed the testing phase with the neurons of the remaining trajectory. For cross-hemispheric validation tasks, we tested the performances of trained right-hemisphere and left-hemisphere models using neurons of the opposite hemisphere.

We have started our machine learning procedure with the classical preprocessing steps: filling the missing values, feature scaling, and standardization[46]. Following these preliminary steps, we defined additional feature selection criteria to our initial mutual information-based approach. As mentioned previously, the work of Mosher et al. exploited the principal component definitions for feature reduction. Following the mutual information-based first feature selection, we needed to conduct an additional step in some cases to further remove candidate neural biomarkers. For example, some candidate neural biomarkers showed isotropic behaviour in terms of mutual information in 3D MNI space (see Supplementary Material Figure 4). Isotropic candidate biomarkers are not particularly suitable to investigate the electrophysiological structure of dystonic GPi. Thus, some of the candidate biomarkers passed the mutual information-based feature reduction but decreased the overall performance of 3D localization tasks. To pursue the use of information theory as our main feature selection mechanism, we adopted an entropy-based approach. We constructed a Random Forest[47] model with the information gain[48] metric which is an index that measures the entropy of the candidate neural biomarker for the classification task to define its importance. Using the same model, we calculated the capability of each candidate neural biomarker concerning the amount of entropy it can describe. We defined then a set of subsets of candidate biomarkers including only those with information gain above iteratively increasing threshold values, and for each of these subsets, we computed the localization performance and its robustness (see Supplementary Materials 6.1. for details). We finally selected the subset with the optimal and robust performance. This final step is hence based on the biomarker set performance, rather than on the individual performance of each biomarker.

We repeated this two-level feature selection approach for all tasks in the prediction block to select for each task the optimal set of biomarkers. For the classification process, we have selected Decision Tree[49], Random Forest[47,49], K-Nearest Neighbors[49], Gaussian Process Classifier[49], and Support Vector Machine[49] as our main classifiers. Because of the class imbalance, we assessed classification performance by computing balanced accuracy, weighted AUC (area under the curve) score, and weighted F1 score. In the training and testing phases of our machine learning models, we adopted the Stratified K-Fold Cross Validation[49] procedure to prevent possible biases of classical training and test split approach. In each split of five-folds[50], we applied SMOTE[49] oversampling with additive white gaussian noise (AWGN) (standard deviation = 0.005) for overcoming imbalanced data problems in the training set. In each fold, we calculated our performance metrics with the test set and averaged scores across all five folds to acquire the mean performance. As a final step in this module, we compared the performances of all five neural decoders across all tasks in the prediction block and selected the best two classifiers for the Voting classifier with a soft voting approach. With the Voting classifier, we tried to further improve the localization performances by generating our unique ensemble model. Instead of using all five trained models, we only selected the best performing two models because, for some classification tasks, we experienced degraded performances in some of the trained classifiers. Hence, including these classifiers in Voting Classifiers reduced the overall performance.

## Results

### 3.1 Feature Selection

Before presenting the results of our two-step feature selection procedure, we wanted to emphasize an important fact. The fraction of neurons displaying oscillatory behaviour is quite low (see Table 1). Since neural oscillations are not significantly evident for dystonic GPi neurons in MERs at the single unit activity level, we did not come across any oscillation-related candidate neural biomarkers to pass the feature selection steps and to be included in the neural decoder in any type of localization.

**Table 1.**
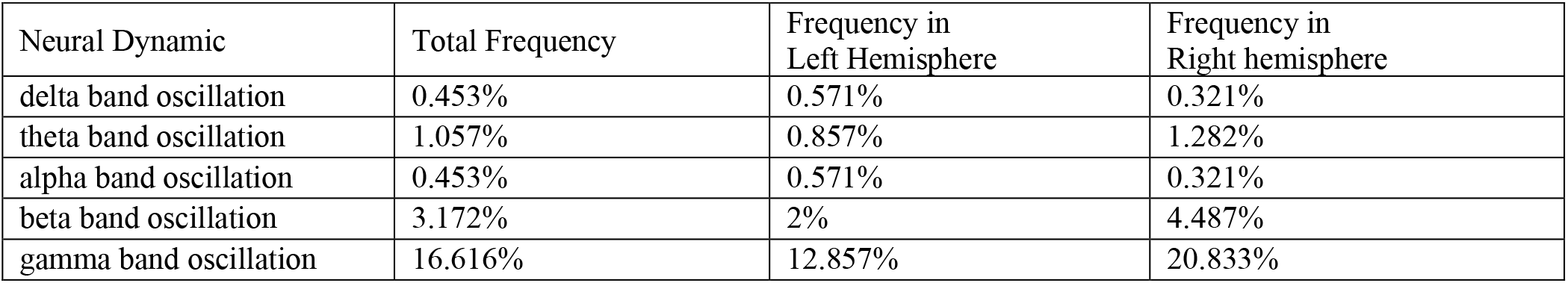
The oscillatory behaviour of single unit activities. The existence of significant neural oscillations in dystonic GPi neurons are quite low. Only the gamma band oscillations are comparably more common with slightly higher than 15%.

| Neural Dynamic | Total Frequency | Frequency in Left Hemisphere | Frequency in Right hemisphere |
| --- | --- | --- | --- |
| delta band oscillation | 0.453% | 0.571% | 0.321% |
| theta band oscillation | 1.057% | 0.857% | 1.282% |
| alpha band oscillation | 0.453% | 0.571% | 0.321% |
| beta band oscillation | 3.172% | 2% | 4.487% |
| gamma band oscillation | 16.616% | 12.857% | 20.833% |

We employed the information theory paradigm to generate significant neural biomarker sets from all candidate neural biomarkers, as indicated in the Methods section. Hence, we were able to achieve a cohesive two feature selection steps with exploding two different metrics: mutual information and information gain in the first and second steps respectively to boost neural decoder performances.

Following our feature selection process in the GPi in-out localization task in the 1D reference frame, we ended up with seven significant neural biomarkers: two firing regularity-related and five power spectral density-related neural biomarkers (green rows in Table 2).

**Table 2.**
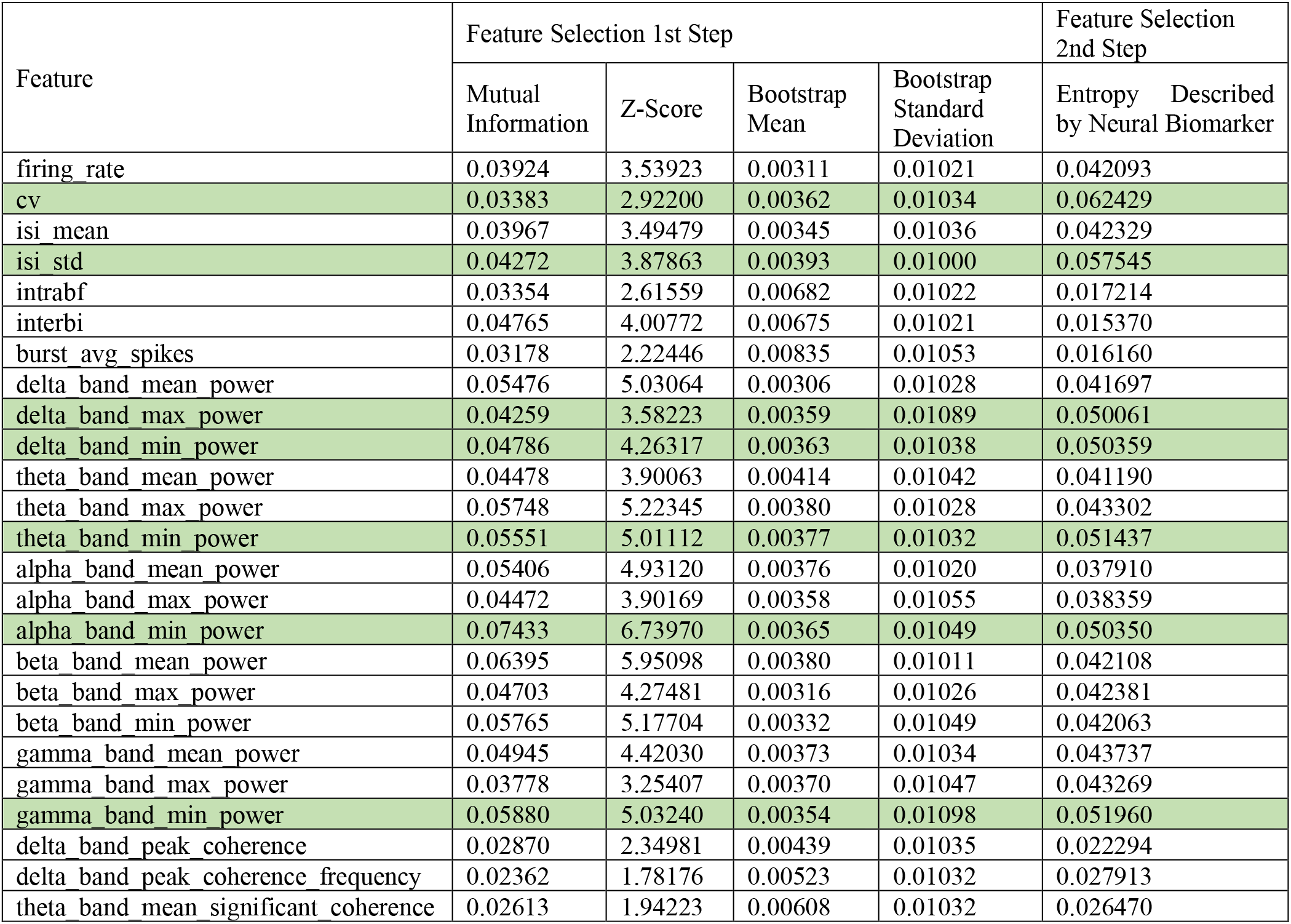
Two-step neural biomarker selection results of GPi in-out neuron localization for merged hemispheres task in the 1D relative depth reference frame. The table presents the results of the feature selection process of GPi in-out localization in 1D reference frame. Only features that show significant mutual information in the first step of feature selection are presented in this table. Green highlighted rows indicate biomarkers selected by the process for this task.

Contrary to other localization tasks, the mutual information-based feature selection step did not eliminate any candidate neural biomarkers for GPi in-out localization in the 3D MNI reference frame. The other analysis scenarios occurred mainly in one direction independently from the reference frames. But for the GPi in-out task in the MNI reference frame, we were considering the 3D structure of the GPi in all directions. Since we were only interested in whether the neuron is located inside GPi, determining mutual information in a specific axis loses relevancy. The other main difference is that we decreased our mean entropy threshold from 0.05 to 0.03 (again a heuristic threshold) (see Supplementary Material Section 6.1.) considering the higher number (no elimination of features previously) of markers evaluated for mean explained entropy. Hence, it got harder for a biomarker to explain the fraction of total entropy by 0.05 individually. We defined two final sets of significant neural biomarkers. The correlation coefficient of ISI distribution, the proportion of spike lies in bursting intervals, the maximum power in the alpha band, the mean and peak coherence in the gamma band was selected for the GPi in-out localization in the left hemisphere. For the right hemisphere, firing regularity, local variation, ISI distribution skewness, the significant portion of mean coherence in the delta band, and mean coherences delta, theta, and alpha frequency bands provided entropy decrease that surpasses the predefined threshold value (see Figure 3D, Supplementary Material Table 8).

In terms of dorsoventral localization in the 3D MNI reference frame, we identified significant neural markers carrying information about the spatial structure of dystonic GPi (see Method, Figure 3E and Figure 4A) for all three tasks in the prediction block. Oscillation-related neural biomarkers were not informative at all in the dorsoventral direction with no biomarker passing the 10% threshold in the first step of our feature selection. In terms of bursting-related markers, 4 out of 6 of them carried some level of information in the dorsoventral axis, but they failed to decrease the entropy of the dorsoventral localization problem adequately (see Supplementary Material Table 9). Some firing rate and regularity-related biomarkers (mean firing rate, the standard deviation of ISI, and coefficient of variation) were generally essential for dorsoventral localization tasks in the prediction block (see Methods and Figure 7B). ISI mean was also labelled as a descriptive marker for the dorsoventral localization tasks in merged and right hemispheres cases. Moreover, we found firing rate, ISI mean and ISI standard deviation for merged hemispheres, cv and ISI standard deviation for left hemisphere GPi were significantly different between dorsal and ventral neurons (the median split: -3.880 on the Z-axis of GPi definition in DISTAL Atlas, p<0.05 Holm-Bonferroni correction applied for Mann-Whitney U test after Cramer-Von Mises test, see Supplementary Material Table 18 and Figure 7A).

**Figure 4.**
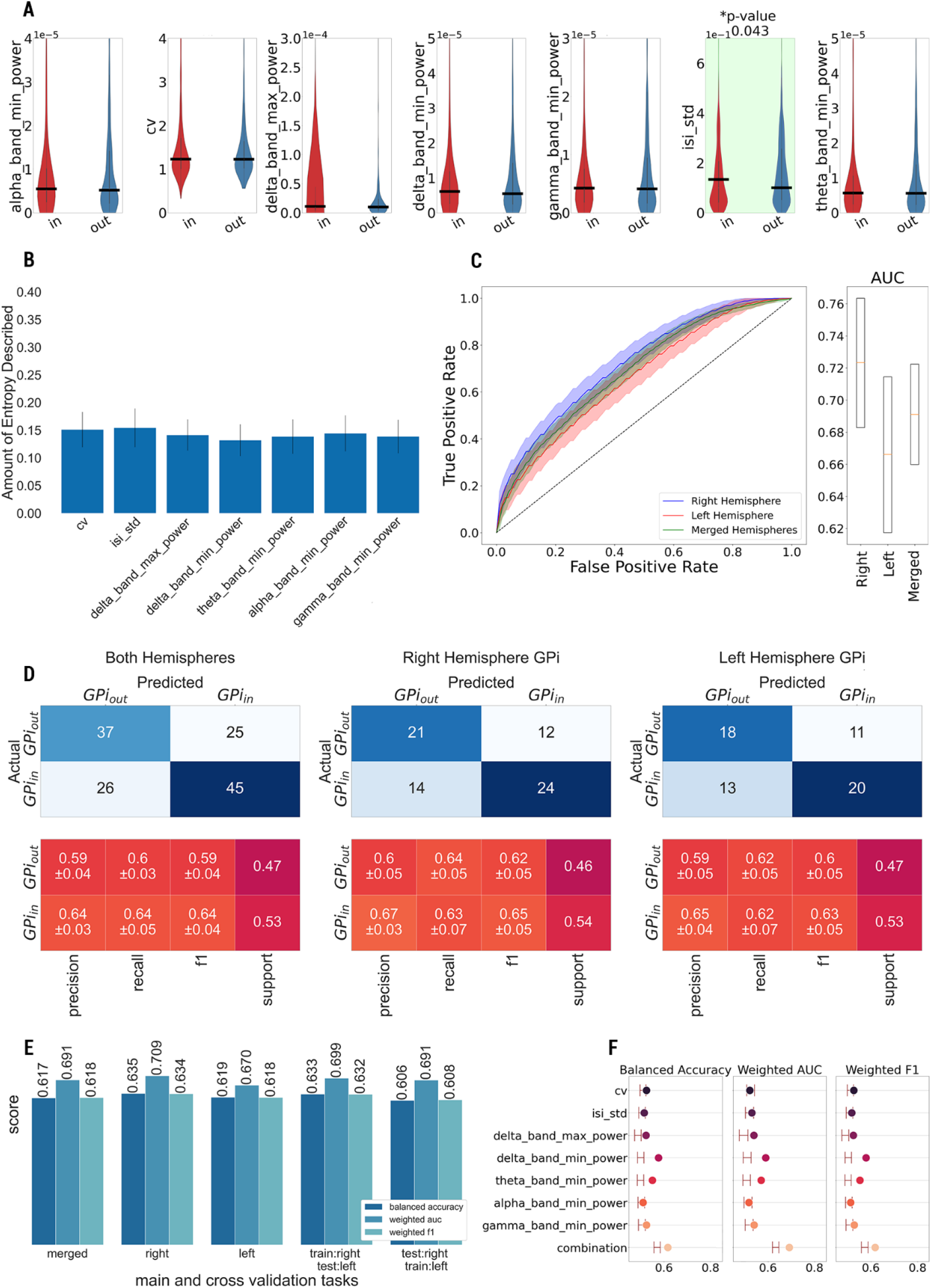
The results of GPi in-out neuron localization in the 1D relative depth reference frame. **(A)** The value distribution of significant neural biomarkers for GPi-in and GPi-out neural populations and their corresponding probability values (corrected with Holm-Bonferroni Method) of Mann-Whitney U tests. **(B)** Significance of neural biomarkers in terms of their abilities to explain mean entropy during the generation of Random Forest Trees classifier (The values are computed again after the completion of the feature selection process with only significant neural biomarkers). **(C)** The ROC curves of the Voting Classifiers (constructed with trained Random Forest Trees and KNN classifiers) for different tasks of prediction block with their corresponding percentile-based confidence intervals (95%) of AUC scores. **(D)** The confusion matrices of GPi in-out classification tasks in right-hemisphere-only, left-hemisphere-only, and merged-hemispheres tasks and their classification performances (precision, recall, and F1 scores). **(E)** The performances of the Voting Classifier models in respect of balanced accuracy, weighted AUC, and weighted F1 scores in tasks of prediction and validation blocks. **(F)** The performance of the Voting Classifier model for single and combined versions of the significant neural biomarkers in merged hemisphere tasks. The percentile-based confidence interval of the random chance scenario for each performance metric is indicated with two horizontal lines (left whisker: lower bound, right whisker: upper bound of confidence interval).

### 3.2 GPi in-out Localization in 1D Reference Frame

In the first classification task, we aimed to localize neurons in terms of their positioning whether inside or outside of GPi in the 1D reference frame across trajectories (see Methods). Based on the intraoperative notes of a neurophysiological expert, we labelled the neurons on trajectories in terms of their location. GPi-in neurons are defined as neurons that are positioned between -4mm and - 10mm from the target location. The remaining neurons were considered to be outside of the GPi.

MERs are performed during DBS implant to support the localization of the target area after the initial guess on its position is performed with pre-surgical imaging. Hence, before moving to use MER data to analyse the difference in the activity over the volume of the GPi, we assessed which of the neural markers (see Methods) could carry more information about the position of a given recording site within or outside the GPi.

Following our feature selection process, we ended up with seven significant neural biomarkers: two firing regularity and five power spectral density-related neural biomarkers (green rows in Table 2). The standard deviation of the ISI distribution was the only neural biomarker significantly differentiated inside and outside the GPi (p<0.05, Mann-Whitney U test, with Holm-Bonferroni correction) (see Figure 4A, Supplementary Material Table 10). However, in terms of the amount of entropy they can explain, neural biomarkers are relatively similar (see Figure 4B). Individual performances of trained machine learning models were evaluated with the Stratified 5-Fold cross-validation with training set upsampling (see Methods). The best performance was achieved with the KNN classifier with 0.623 mean balanced accuracy, 0.692 mean weighted AUC score, and 0.624 mean weighted F1 scores across all five folds. Random Forest Tree algorithm, the second-best performing model, provided a slightly worse performance with a 0.619 mean weighted AUC score. We used both KNN and Random Forest models in an ensemble model to generate a Voting Classifier with the soft voting mechanism (Supplementary Material Table 11). To measure the robustness of the Voting Classifier, we applied a bootstrap test with 200 iterations (each iteration has also Stratified 5-Fold cross-validation). We acquired the ROC curves across all iterations and generated the mean ROC curve with the 95% percentile confidence interval around it which is given in Figure 4C. This classification was performed for each hemisphere separately and then for the merged hemispheres. Based on the mean ROC curve, the Voting classifier trained for left hemisphere GPi has the best performance followed by merged hemisphere and right-hemisphere classifiers. Apart from the ROC curves, we also illustrated the mean AUC scores and their corresponding confidence interval with 95% percentiles in the right panel of Figure 4C, and the performance differences are also evident in terms of AUC scores, although with similar ranges for confidence intervals of mean AUC score. Note that the class imbalance is not a big problem for GPi in-out classification in the 1D reference frame since we have almost 47% of neurons situated inside of the GPi and 53% remaining located outside of the GPi (see Figure 4D). The performance similarities across all tasks in prediction and validation blocks are also notable in this type of localization. The mean precision, recall, and accuracy scores range in a limited region from 0.59 to 0.65 across all prediction block tasks. Mean balanced accuracy, mean weighted AUC score, and mean weighted F1 scores of all tasks in prediction and two cross-hemispheric validation tasks are given in Figure 4E. In all tasks in the prediction block, we observed approximately 0.7 mean weighted AUC scores. The remaining two performance metrics were also quite similar. It is notable that, even though we trained Voting classifiers with the neurons of one hemisphere and test them with the neurons of the opposite hemisphere, the performance is again similar. Hence, the hemispheric effect for dystonic GPi to decide whether the neuron is inside or outside of the GPi is not crucial for the 1D reference frame.

Further, we assessed the localization capabilities of selected neural biomarkers individually by comparing them to the combined version, the ideal neural decoder. To achieve this final analysis, we trained the previously generated Voting Classifiers with only a single neural biomarker and computed the mean of performance metrics across five stratified folds, with the same bootstrap (N=200) estimation of the performance described above. Individual performances of the coefficient of variation, the standard deviation of ISI distribution, and the minimum power in alpha and beta bands occurred within the random chance intervals (Figure 4F). The minimum power in the delta band showed the best individual performances across all seven selected neural biomarkers. No neural biomarkers delivered a higher or comparable result similar to the combined version of the biomarkers. Overall, the results were robust and significant, but the performance was far from satisfying and required the combination of multiple markers. We concluded that the ability of MER in identifying GPi boundaries by performing the analysis in the 1D space given by penetration tracks is limited in our dataset.

### 3.3 GPi in-out Localization in 3D MNI Reference Frame

To localize the neurons based on the 3D boundaries of GPi, we resorted to the 3D distribution of firing properties in MNI space (see Methods). Briefly, we designed a similar neuron localization analysis in MNI space considering the anatomical structure of GPi using the DISTAL Atlas. To establish the borders of GPI in the MNI reference frame, we used a 0.5 probabilistic value as our heuristic threshold for the DISTAL atlas. This section will provide the complete results of the individual hemispheric tasks in the prediction block of this localization task. Indeed, the performance of GPi in-out localization is significantly decreased by merging the hemispheres (see Supplementary Material Table 15).

Initially, we labelled all neurons as GPi-in or GPi-out in both hemispheres. As previously indicated, we leverage the same computational pipeline for all localization tasks to have comparable results. We initiated our second localization task with feature selection again.

Only, the maximum power in the alpha band significantly differentiated in left hemisphere GPi (p-value = 0.0011 after the Mann-Whitney U test with Holm-Bonferroni correction) (see Supplementary Material Table 14, Figure 5A, the top panel). Contrary to the previous localization task, we did not have a uniform entropy behaviour across all markers. The entropy behaviour of seven markers in the right hemisphere was considerably uniform with a value of 0.15. Contradictorily, we had a more uneven explained entropy distribution for the left hemisphere (see Figure 5B). As mentioned above, we had huge performance differences between merged and split hemisphere tasks in the prediction block. The mean weighted AUC scores reached 0.78 for both the right and left hemisphere GPi, meanwhile, it stayed at 0.631 in the merged hemisphere case. The performance of the trained neural decoder further decreased in the cross-hemispheric cross-validation tasks reaching below 0.62 mean weighted AUC score (see Figure 6B). Similar to selected performance metrics, the behaviour of acquired ROC curves in the bootstrap test with 200 iterations shows parallelism. We acquired mean AUC scores of 0.773 and 0.792 for the right and left hemispheres respectively (see Figure 5C, the right panel). Even the upper limit of the 95% confidence interval of the merged hemisphere neural decoder was not able to surpass 0.70 AUC scores. This situation can be explained for multiple reasons. Initially, when we tried to measure population-wise median differences, since none of the neural biomarkers follows the normal distribution, no marker showed a statistically significant difference between the two neural populations in the merged-hemisphere task. We further investigated to reveal the root cause of performance degradation as follows: we compared biomarkers for GPi-in and GPi-out populations of opposite hemispheres to see if we had any significant differences. Surprisingly, power spectral density-related fifteen biomarkers differed significantly between GPi-in and GPi-out populations of both hemispheres (alpha=0.05). Additionally, minimum power in the gamma band, peak coherence in the alpha band, and peak coherence in the gamma band were the three markers that showed significant population median differences for both GPi-in and GPi-out populations between the two hemispheres (see Supplementary Material). The third possible reason could be related to the drastically different class distribution in both hemispheres: 70(20%) GPi-in, 280(80%) GPi-out in the right hemisphere, 254(81%) GPi-in, and 54(19%) GPi-out neurons. Notably, the class distribution was exactly the opposite in both hemispheres. Even though the merger of hemispheres for complete analysis had an almost balanced class distribution (49% GPi-in, %51 GPi-out), it did not provide better localization performance. The performance superiority of split hemisphere scenarios also was supported by the capabilities of classifiers to label majority class neurons correctly with moderate success in the minority neuron classes (see Figure 6A).

**Figure 5.**
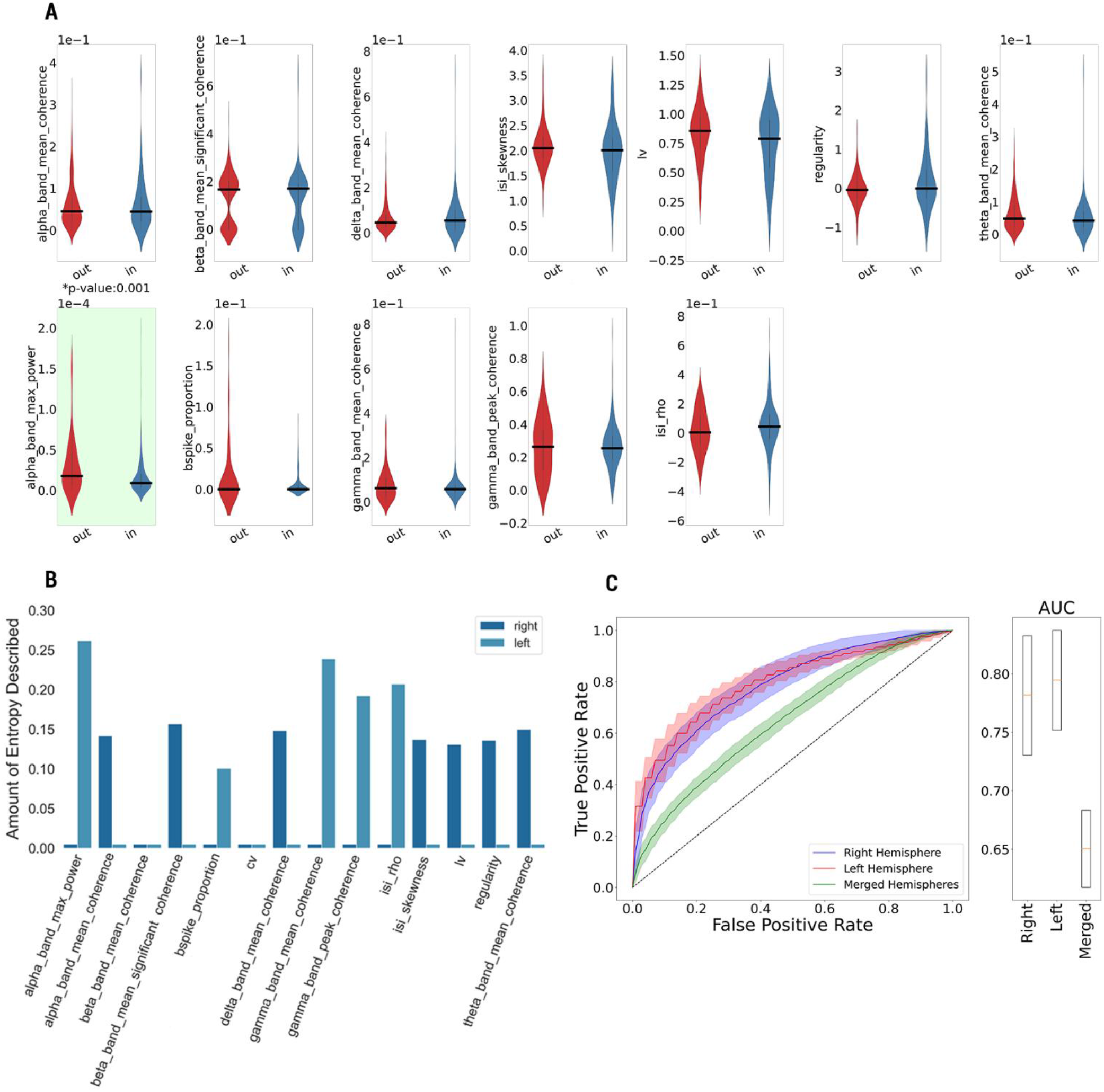
The results of GPi in-out neuron localization in the 3D MNI reference frame. **(A)** The value distribution of significant neural biomarkers for GPi-in and GPi-out neural populations and the corresponding probability values (corrected with Holm-Bonferroni Method) of Mann-Whitney U tests (top: right hemisphere neurons, bottom: left hemisphere neurons). **(B)** Significance of neural biomarkers in terms of their abilities to explain mean entropy during the generation of Random Forest Trees classifier for right hemisphere and left hemisphere neurons individually. **(C)** The ROC curves of the Voting Classifiers (constructed with trained Random Forest Trees and Gaussian Process classifiers) for different analysis scenarios with their corresponding percentile-based confidence intervals of AUC scores.

**Figure 6.**
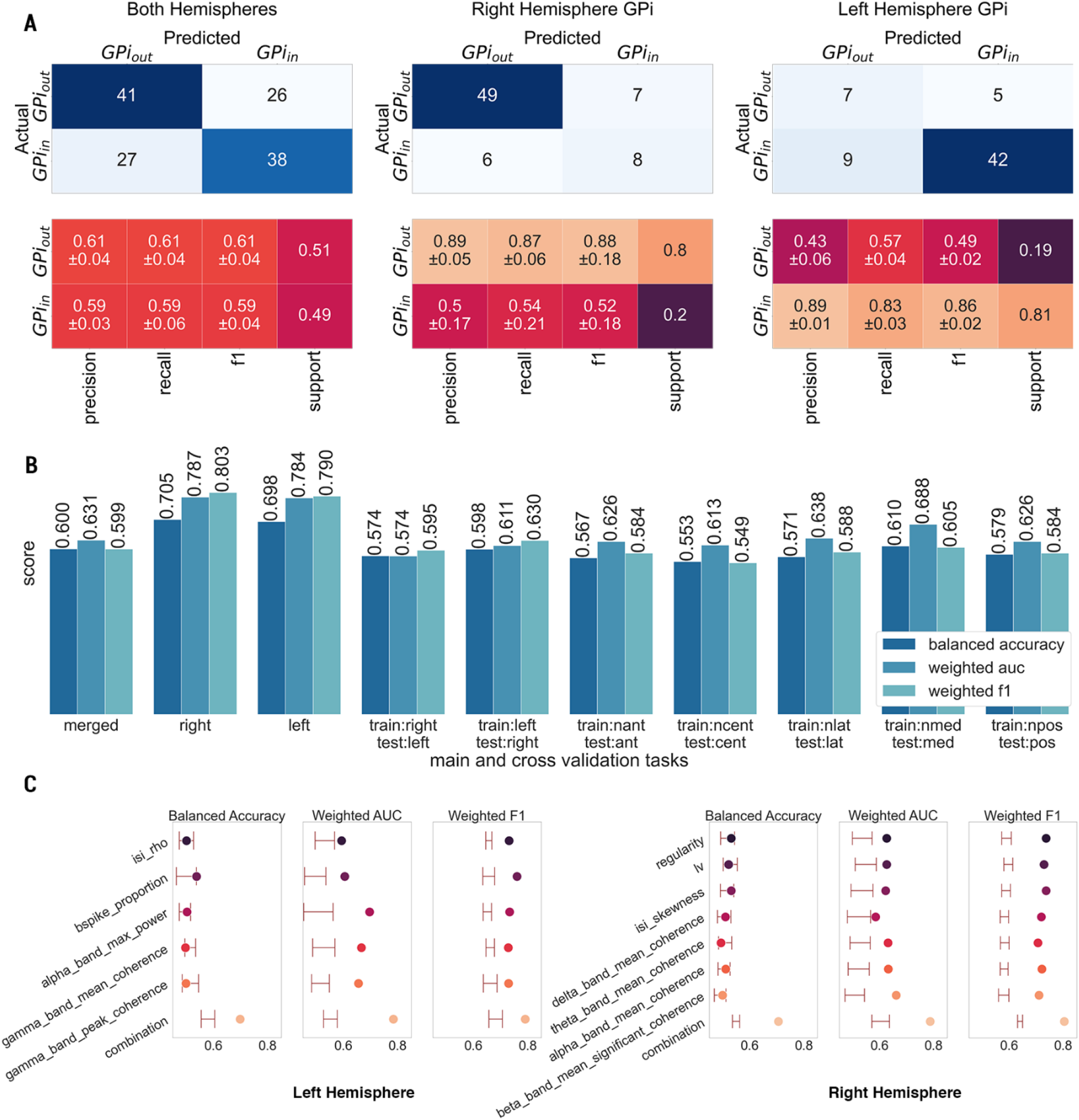
The results of GPi in-out neuron localization in the 3D MNI space domain. **(A)** The confusion matrices of GPi in-out classification tasks in right-hemisphere-only, left-hemisphere-only, and merged-hemispheres analysis modes and their classification performances (precision, recall, and F1 scores) in MNI space. **(B)** Performance of the Voting Classifier models using the balanced accuracy, weighted AUC, and weighted F1 scores in tasks of prediction and validation blocks. (trajectories: and(anterior), cent(central), lat(lateral), med(medial), pos(posterior), nant(non-anterior), ncent(non-central), nlat(non-lateral), nmed(non-medial), npos(non-posterior). **(C)** The performance of the Voting Classifier model for single and combined versions of the significant neural biomarkers in merged hemisphere analysis scenario. The percentile-based confidence interval of the random chance scenario for each performance metric is indicated with two horizontal lines (left whisker: lower bound, right whisker: upper bound of confidence interval).

In terms of individual capabilities of biomarkers in both hemispheres, they were not able to provide a better mean balanced accuracy performance than the 95% confidence interval of random chance (see Figure 6C). The high F1 scores of individual markers and confidence interval of random chances that were constructed with F1 are mostly the side effect of keeping the class distribution during the random chance bootstrap test. In the worst-case scenario, 60% of majority class neurons will be labelled the same since the minority neural population is only made up of 20% of all neurons in both hemispheres. The power of the combined version of neural biomarkers for both hemispheres was especially more evident in this neural localization task.

### 3.4 Dorsoventral Localization of Spike Dynamics in GPi

We next investigated whether the neural dynamics identified so far differ across the spatial structure of the dystonic GPi (in particular, along the vertical dorsoventral axis). Akin to GPi in-out localization in the 1D reference frame, we will present the complete analysis results of the merged hemisphere task of the prediction block.

The discrimination capabilities of some of the individual markers (the standard deviation of ISI distribution and coefficient of variation) were close to the combined version of biomarkers and way higher than the chance within the 95% confidence range (see Figure 7F). However, the combined version of neural biomarkers for Voting Classifier (KNN with 0.628 mean balanced accuracy, 0.690 mean weighted AUC score, 0.638 mean weighted F1 score, and RF with 0.574 mean balanced accuracy, 0.602 mean weighted AUC score, 0.608 mean weighted F1 score) helped us to define the most robust neural decoder (see Supplementary Material Table 19). Our neural decoder has reached 0.649 mean balanced accuracy, 0.676 mean weighted AUC, and 0.661 mean weighted F1 scores and it performed significantly higher than 95% random chance confidence intervals (0.543, 0.565), (0.599, 0.644), (0.559, 0.583) respectively (see Figure 7F). The ROC curves of the main analysis tasks had a mean AUC score of 0.7. The only difference was the merged hemisphere task had the narrowest confidence interval with 95% confidence lying between 0.649 to 0.738 after 200 bootstrap iterations. (See Figure 7C, the right panel). Meanwhile, the right hemisphere decoder had a broader confidence interval since the right hemisphere drastically lowers the number of neurons located within GPi compared to the left hemispheres (70 and 254 respectively).

**Figure 7.**
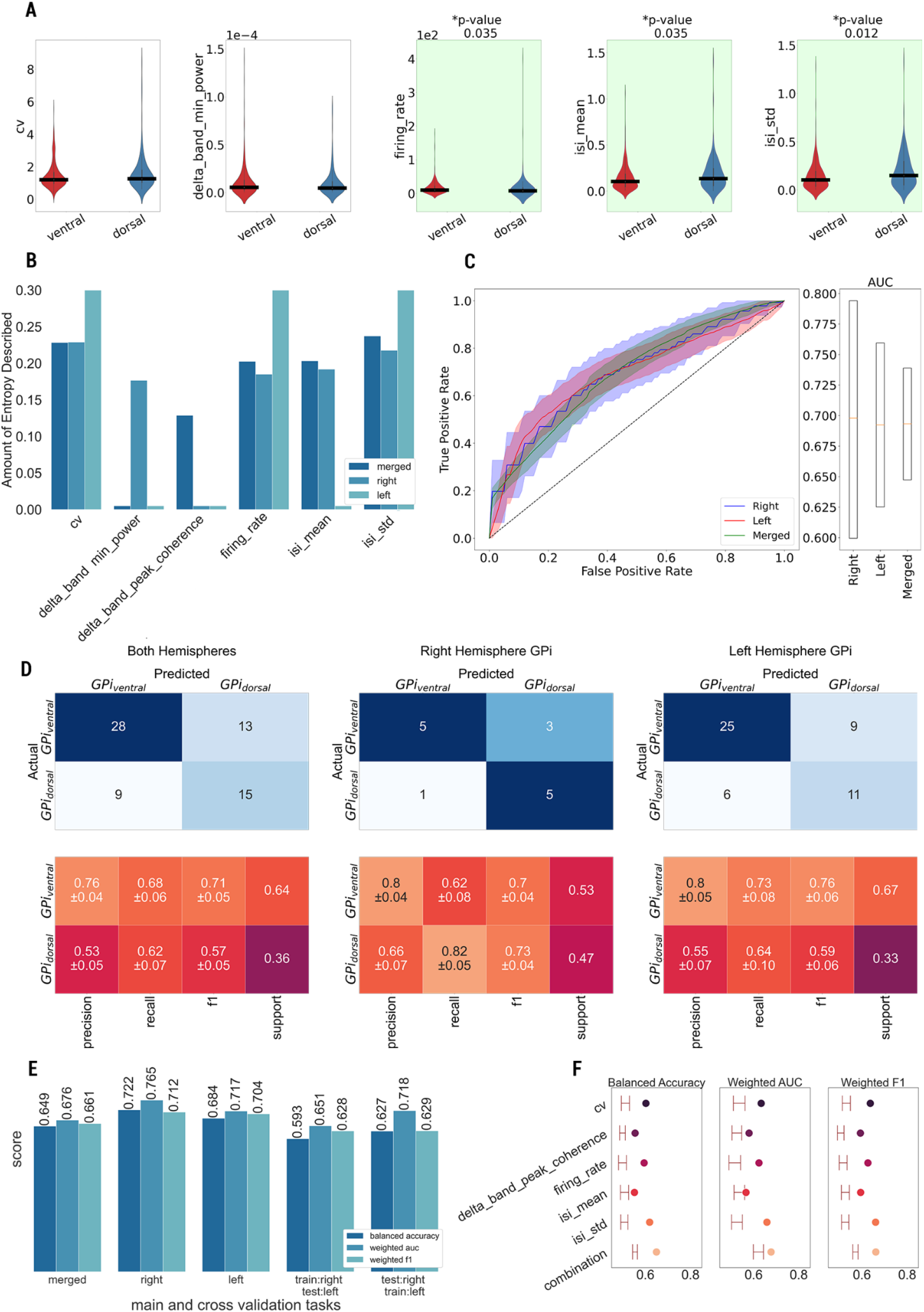
The results of dorsoventral neuron localization within GPi in the 3D MNI space domain. **(A)** The value distribution of significant neural biomarkers for dorsally and ventrally located neural populations and the corresponding probability values (corrected with Holm-Bonferroni Method) of Mann-Whitney U tests in merged hemispheres analysis task. **(B)** Significance of neural biomarkers in terms of their abilities to explain mean entropy during the generation of Random Forest Trees classifier for dorsoventral localization task in all tasks in prediction block. **(C)** The ROC curves of the Voting Classifiers (constructed with trained Random Forest Trees and KNN classifiers) for different analysis scenarios with their corresponding percentile-based confidence intervals of AUC scores. **(D)** The confusion matrices of GPi dorsoventral classification tasks in prediction block tasks and their classification performances (precision, recall, and F1 scores) in MNI space. **(E)** The performances of the Voting Classifier models in respect of balanced accuracy, weighted AUC, and weighted F1 scores in tasks of prediction and validation blocks. **(F)** The performance of the Voting Classifier model for single and combined versions of the significant neural biomarkers in a merged hemisphere analysis scenario. The percentile-based confidence interval of the random chance scenario for each performance metric is indicated with two horizontal lines (left whisker: lower bound, right whisker: upper bound of confidence interval).

The class imbalance was more visible in the confusion matrices of prediction block tasks (see Figure 7D). We had class support for machine learning models 65% and 35% for ventrally and dorsally located neurons in merged hemispheres, and left hemisphere cases. Even though we had a more balanced neuron distribution in the right hemisphere, the low number of neurons created the main challenge in this task. The performance of neural decoders was relatively better when we conducted dorsoventral localization in right and left hemisphere GPi separately by a really small margin. The results of cross-hemispheric cross-validation tasks were comparatively lower than their prediction block counterparts. Especially performance decreased in the cross-validation task when the Voting Classifier is trained with 70 right hemisphere model and tested with 254 left hemisphere GPi neurons over stratified 5-fold cross-validation. The mean balanced accuracy dropped to 0.593 which is the lowest across all tasks in this localization procedure (see Figure 7E).

## Discussion

Our statistical analyses prove that some neural biomarkers related to specific spiking phenomena vary both in 1D relative depths and 3D MNI space. This is in striking contrast with the fact that GPi is considered a homogeneous subcortical structure. The results of this study imply that the variety of neural dynamics inside dystonic GPi is high because we were able to define directions that showed statistically significant changes in terms of some neural biomarker values based on mutual information metrics. We used these results to locate neurons in three different localization tasks where we have comparable high classification performances for locating neurons based on different criteria with selected neural biomarkers. In our study, we try to collaborate on different neural biomarkers that are related to different spiking phenomena of dystonic GPi neurons. We can group these neural biomarkers into the following main categories: firing rate, firing irregularity, bursting, oscillation, spectral coherence, and power spectrum-related neural biomarkers. Even though we have found neural oscillations on MERs at the single neuron level, especially in the gamma band, frequency domain-associated neural biomarkers fail to describe the position of neurons in the dorsoventral localization task where we only consider neurons that are located inside the dystonic GPi. This result can be also interpreted from a neurological point of view. It may imply that the neural behaviour in terms of bursting, oscillation, and other spectral components of neural activity stay homogenous across the GPi at the single neuron level. Thus they are not reliable neural phenomena to locate the neurons inside the GPi. The other important fact, we have some significant frequency domain (oscillation, bursting, or coherence-based) neural biomarker that shows significance in terms of mutual information in the 1D relative depth frame. First and second-order statistics such as firing rate and firing regularity-related neural biomarkers vary significantly inside dystonic GPi.

It is important to note that our approach did not assume any a priori knowledge of the neurophysiological properties of the candidate neural biomarkers, as we only measured each biomarker’s capacity to carry enough information to locate the DBS electrodes algorithmically within GPi. In future studies, we will address the underlying mechanism leading to spatial inhomogeneity of specific features of dystonic GPi activity.

### 4.1 Limitations

As cited earlier, we were not able to corroborate the anatomical position of the MERs electrodes with preoperative or postoperative medical images. Hence, the use of normalized probability-based anatomical atlas for the confirmation does not affirm the inter-subject anatomical variabilities of our subject group. The possibility of brain tissue displacements during GPi DBS surgery further added uncertainty to our findings. Furthermore, we believe that the GPi bounds were set conservatively by setting the heuristic threshold for the DISTAL atlas as 0.5. The number of neurons in the right GPi hemisphere is roughly 20%, and the majority of the electrode trajectories are outside of the right GPi. We know that before craniotomy, patients’ heads are tilted at a precise angle. The tilt could potentially cause these types of misalignment between the real position of neurons and their DISTAL atlas-based location. Even though the use of a normalized atlas introduced some level of limitations, the DISTAL atlas is a widely adopted and validated atlas for DBS studies. In a recent study, MERs and DISTAL atlas were found to be consistent in determining the subthalamic nucleus (STN) entry for Parkinson’s disease patients who underwent STN DBS surgery[18].

The expert definition was used for defining GPi borders in the 1D reference frame, as previously described. The concept introduces a level of uncertainty because we did not have relative locations of electrode trajectories to GPi.

We repeated the neuron localization procedure also relative to the mediolateral and anteroposterior axes. The main issue was we only had neurons located in the anterior and lateral sections of dystonic GPi in both hemispheres due to the location of trajectories of DBS electrodes. Hence, we were not able to discriminate neurons in all anatomical axes to get the complete functional structure of the dystonic GPi.

We are aware that working with SUAs that are isolated from the MER recordings poses methodological difficulties in terms of oscillatory behaviour detection, especially in single-trial experiments. Consequently, we do not assert that neural oscillations are not part of the pathophysiology of dystonia within the GPi. In our analysis, we only concluded that neuronal oscillations are not the optimal feature for lead localization based on MERs in GPi DBS surgeries for dystonia.

Finally, the acquired dataset was sufficient to lead to statistically robust results, mainly due to the high number of tracks per patient and the high quality of the recordings, but the set of patients was however limited to ten. Before translating to clinical application, larger datasets should be analysed to ensure that the proposed methods can overcome inter-subject differences.

### 4.2 Algorithms performance

As we suggested in our introduction, the peculiar characteristics of the transition from the external segment of the globus pallidus (GPe) to GPi, such as the sparseness of the neurons along the pallidum, the absence of a marked reduction in neuronal activity, and the high discharge rate variability, challenged the electrophysiological determination of GPi borders and led to AUC values of 0.67. Additionally, there was no well-established anatomical delineation between the dorsal and ventral regions of GPi for the dorsoventral location. As a result, we divided the GPi using the median split: -3.880 on the GPi Z-axis, which could also contribute to the performance of our pipeline. Hence, moderate values of AUC are less an indication of the efficacy of the method than a measure of the structural inhomogeneity of GPi. Apart from the neuroanatomical aspect of the GPi and the surrounding tissue, several factors (the issue regarding postoperative imaging, and conservatively defined GPi boundaries based on the Distal Atlas that we mentioned in our Limitation Section) introduced this uncertainty for the neuron location localization procedure, again leading to an AUC in the order of 0.67. However, cross-validation analyses and confidence interval estimation confirmed the robustness of the classification procedures.

We experimented with different classification approaches that exploit several mathematical concepts to distinguish particular neuron populations. Initially, we employed two tree-based classification algorithms: Decision Tree Classifier and Random Forest Classifiers. Both algorithms construct tree structures based on the capability of neural biomarkers to reduce impurity during the classification. The main advantage of the Random Forest Classifier is generating several random Decision Trees and conducting training instantaneously. The outcome of the classification process will be determined based on major votes between different trees. Being an ensemble approach, it generally provides robust results. For the K-Neighbour Classifier, the spatial structure of the dataset plays a primary role; definition of the distance metric, we used Euclidean distance, and the neighbour radius criteria can affect the classification results drastically. Considering that neural dynamics are entirely complex, the definition of apparent borders between neuron populations is not achievable for all localization procedures. Correspondingly, the spatial structure is vital for the classification of Support Vector Machines. Since SVM utilizes the definition of hyperplanes that separates the regions that belong to different classes, not all problems have a suitable structure for SVM. In the Gaussian Classifier case, the algorithm attempts to exploit the Gaussian probability distribution function similarly to Gaussian mixture problems. It tries to estimate the Gaussian distribution of each neural biomarker for each class in terms of mean and variance properties. Additional to these parameters, gaussian processes require specifying a kernel that manages how neurons relate to each other; specifically, it defines the covariance function of the data. The main problem might occur when the distributions of two classes for a specific neural biomarker are noticeably similar. In this case, the algorithm cannot be able to discriminate neuron populations.

### 4.3 Lateralization

Our results posit that dystonic GPi may have a potential lateralization effect in terms of the in-out localization task conducted in the MNI reference frame. As stated in the result section, no neural biomarkers were selected for the neural decoders that were constructed for the right and left hemispheres individually. Furthermore, merging the neurons of two hemispheres onto the right GPi resulted in significant performance degradation.

The lateralization effect could be originated from two different sources. Firstly, the craniotomy for GPi DBS surgeries is initiated from the right side of the cranium of dystonic patients. Besides, when we examined the baseline motor indexes of the Burke-Fahn-Marsden Dystonia Rating Scale (BFMDRS) for the right arm, left arm, right leg, and left leg of patients, we discovered that the right sides of half of the patients are more affected by primary dystonia compared to their left sides (remaining 30% equally affected and 20% no baseline scores) (see Supplementary Material Table 22).

On the contrary, electrophysiological dynamics within GPi are consistent for two hemispheres, and the primary dystonia mostly modulates the mean firing rate and firing regularity of neurons.

### 4.4 Perspectives

We developed a method able to describe the functional structure of the GPi based on intraoperative multichannel high-density MERs. This approach can be fruitfully applied to find disease-specific and patient-specific partitioning on the GPi and hence support the localization of optimal DBS targets. Crucially, this method could also be applied to other neurological conditions to be treated by GPi implant, i.e., Tourette’s syndrome and Parkinson’s disease.

The proposed computational pipeline has the feasibility to operate in an online manner during DBS surgeries for lead localization. With a few straightforward changes to the pipeline, offline-to-online translation may be achieved. As an initial step, one of the recently proposed high-performance online spike sorting algorithms[51–53] can replace the offline counterpart that we utilized in our pipeline. Since each spike sorting algorithm adopts different computational and statistical approaches, the SUA generation time will differ. We approximated the computational running time of these online algorithms for 10-second recordings based on their reported benchmark tests. Mostly, they can extract single units in less than 1.5 seconds (starting from 0.64 to 1.49 seconds). Approximately 1.25±0.24 (follows the Gaussian distribution for 100 neurons) seconds are needed to produce all the neural biomarkers we presented in the study for each neuron. Considering that the voting classifier was previously trained before the surgery, the localization of the neuron by the trained neural decoder takes around 0.98 seconds. With the aforementioned modifications, we thus think that an online implementation of our computational pipeline can predict the location of DBS leads in around 3.68 seconds (including the online spike sorting procedure) for each 10-second MER recording. In our dataset, we had an average of 10 depth levels per trajectory across 10 patients. Considering this fact, the total running time of the online version of the proposed pipeline takes around 37 seconds during the whole MER collection procedure for each trajectory. Additionally, the value distribution of significant neural biomarkers may also be illustrated, and the lead localization can be displayed on an anatomical atlas in real time. As a result, the pipeline we demonstrate here may serve as the basis for software that takes advantage of all the features and low computational time for providing feedback to neurosurgical teams in real-time.

## Data Availability

All data produced in the present study are available upon reasonable request to the authors

## Data Availability

Scientists can obtain access to individual-level data from the UK Biobank by applying to UK Biobank (https://www.ukbiobank.ac.uk/enable-your-research).

## Ethical Approval

The study was approved by the Carlo Besta Institutional Ethics Committee (reference number 7/2014) and was carried out according to the Declaration of Helsinki. Informed written consent was obtained either from the patients or from their legal representative.

