## Supplementary Material for "Mapping the electrophysiological structure of dystonic Globus Pallidus pars interna through intraoperative microelectrode recordings"

### 1. Patient Demographic

| Patient ID | Follow-Up Period (Months) | Genetic Factor | Classification | Age Range at Implantation | Disease Duration | Age Range at Onset | Baseline Motor Score | Baseline Disability Score |
|---|---|---|---|---|---|---|---|---|
| 1 | 84 | DYT1 | Segmental | 41-45 | 19 | 11-15 | 10 | 2 |
| 2 | 12 | DYT1 | Segmental | 51-55 | 25 | 31-35 | 24 | 3 |
| 3 | 48 | None | General | 46-50 | 15 | 21-25 | 84 | 17 |
| 4 | 12 | DYT1 | General | 31-35 | 26 | 16-20 | 60.5 | 17 |
| 5 | 6 | None | Multifocal | 61-65 | 60 | 1-5 | None | None |
| 6 | 96 | None | Segmental | 31-35 | 16 | 11-15 | 24 | 3 |
| 7 | 12 | DYT1 | Multifocal | 41-45 | 37 | 6-10 | None | None |
| 8 | 120 | DYT1 | General | 46-50 | 21 | 6-10 | 31.5 | 18 |
| 9 | 132 | DYT1 | Segmental | 61-65 | 46 | 16-20 | 29 | 11 |
| 10 | 48 | DYT1 | Segmental | 41-45 | 13 | 26-30 | 12 | 3 |
| **Total** | 57 ± 45.47 | 7Y/3N | | | 27.8 ± 15.31 | | 34.37 ± 25.33 | 9.25 ± 7.27 |

**Table 1 - Clinical and Demographic Information of Study Group**
The table shows the demographical and clinical information regarding GPi-DBS surgery. In total, 10 dystonic patients are involved in the scope of the project.

### 2. Microelectrode Recordings Acquisition

| | Left Hemisphere | | | | | Right Hemisphere | | | | | Depth | |
|---|---|---|---|---|---|---|---|---|---|---|---|---|
| **Patient ID** | **A** | **C** | **L** | **M** | **P** | **A** | **C** | **L** | **M** | **P** | **Min** | **Max** |
| **1** | 13 | 13 | 0 | 13 | 13 | 13 | 13 | 13 | 13 | 13 | -10 | +1 |
| **2** | 22 | 22 | 22 | 22 | 22 | 20 | 19 | 17 | 20 | 20 | -10 | +3.5 |
| **3** | 20 | 20 | 20 | 20 | 20 | 15 | 15 | 15 | 15 | 15 | -10 | +2 |
| **4** | 20 | 4 | 0 | 0 | 0 | 20 | 20 | 0 | 20 | 20 | -10 | +1 |
| **5** | 13 | 16 | 18 | 18 | 18 | 17 | 17 | 0 | 18 | 0 | -10 | +2 |
| **6** | 14 | 14 | 14 | 14 | 14 | 17 | 17 | 17 | 0 | 17 | -10 | +2 |
| **7** | 22 | 22 | 22 | 22 | 22 | 20 | 20 | 0 | 20 | 20 | -10 | +3 |
| **8** | 21 | 21 | 21 | 21 | 21 | 28 | 28 | 28 | 29 | 28 | -10 | +1 |
| **9** | 5 | 5 | 2 | 6 | 13 | 0 | 17 | 17 | 17 | 17 | -9 | +1 |
| **10** | 22 | 22 | 22 | 22 | 22 | 24 | 24 | 24 | 24 | 24 | -10 | +2 |
| **Population Level** | 172 | 159 | 141 | 158 | 165 | 174 | 190 | 131 | 176 | 174 | **1640** | |

**Table 2 - MER Distribution by GPi Tracks**
The distribution of MER recordings across depths of the electrode trajectories for all patients. The red cells denoted with 0 represent the tracks where there is no MER recording exists.

| The number of recordings per depth | Count | Percentage |
|---|---|---|
| One Recording | 690 | 63,24 |
| Two Recordings | 295 | 27,04 |
| Three Recordings | 78 | 7,15 |
| Four Recordings | 17 | 1,56 |
| Five Recordings | 11 | 1,01 |
| | 1091 | 100 |

**Table 3 - MER Distribution by Number of Recordings in Tracks**
The distribution of the number of MER recordings per track. In our study, 1091 different MER recordings are collected from 10 dystonic patients. For most of the depth, single MER recordings are available at around 63,24%

For some trajectories, multiple recordings were acquired from the same depth level in a subsequent trend. For example, 11 depth tracks in the whole dataset have 5 electrophysiological recordings, which correspond to 1,01% of all recordings. The track term represents a depth level in a specific trajectory of one of the hemispheres. In this study, there were 1091 individual tracks exist. Detailed information can be found in Table 2.

#### 2.1. MER Configuration

In both hemispheres, the MER trajectories are parallel to the GPi's dorsoventral axis, with the central trajectory crossing the DBS target. As is evident, DBS targets completely match the center trajectories in the mediolateral-anteroposterior plane of GPi in both hemispheres. DBS targets are located within the GPi according to the in-out GPi criterion that we used for the 1D reference frame (Figure 1). For the 3D reference frame, all the trajectories for 8/10 subjects pass through GPi (for the others, anterior and lateral tracks are situated outside) in the left hemisphere. 2/3 of the depth levels of these penetrating trajectories are located inside the GPi based on the 3D definition of the GPi. In terms of the right hemisphere, 6 medial, 3 central, and 7 posterior trajectories of 10 subjects penetrate the GPi (again, based on the DISTAL atlas definition). Similar to the left hemisphere, again, 2/3 of the depth levels coincide within the GPi.

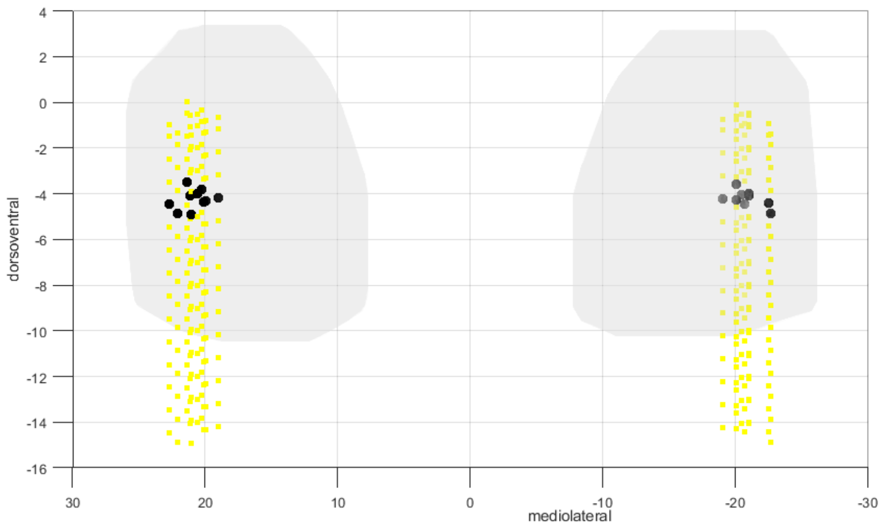


**Figure 1 - The configuration of MERs central trajectories and DBS targets in the 3D MNI reference frame**
The central trajectories of each patient for the right and left hemispheres are in yellow (the same colour convention used in Figure 1A in the manuscript). DBS target positions are represented with black dots.

### 3. Data Preprocessing

In this section, all preprocessing methods applied to the raw data shall be described in all detail. Preprocessing of neural recordings is an essential part of further statistical analyses. The rationale behind preprocessing in this study is transforming the raw data into convenient features, which can be used for neural biomarker generation.

#### 3.1. The merger of Multiple MERs in the Same Track

As mentioned above, there are multiple consecutive MER recording exists for some tracks. This type of segmentation on the raw MER recording splits the data into different epochs. Following this division, each epoch is examined for the existence of artefacts. In our dataset, each epoch represents a section of 10 seconds in the continuous MER recording. Different recordings in the same depth level within a trajectory are merged based on a widely used variance coefficient[2] as the stationarity criterion. This criterion is used to decide whether the recordings need to be sorted individually or together. Invariance equation $x_{k+i}[i]$ indicates the value of k+i spike in spike train and $\bar{x}_{k+i}$ represents the mean value.

$$Variance_{coef} = \frac{\frac{1}{N}\sum_{n=0}^{N}(x_{k+i}[i] - \bar{x}_{k+i})^2}{\frac{1}{M}\sum_{n=0}^{M}(x_k[i] - \bar{x}_k)^2}$$

$$Result = \begin{Bmatrix} Variance_{coef} \leq 2, & merge \\ Variance_{coef} > 2, & split \end{Bmatrix}$$

A critical threshold value for the variance coefficient is defined as 2. If the subsequent epoch of recording in a depth has a variance coefficient higher than 2, these recordings are evaluated individually as two separate data. If the variance coefficient is lower than 2, the merged version of the two recordings shall be analyzed together in the following steps.

#### 3.2. Spike Sorting

In principle, the spikes fired by a neuron recorded by a given electrode have a distinct shape that depends on the morphology of its dendritic tree and the distance and orientation relative to the recording site, among other factors[3]. Spike sorting algorithms applies wavelet transformation to extract a set of features. Then these features are used for clustering all observed spikes into different groups, which represent the activities of single neurons[4]. In general, spike sorting algorithms have mainly four steps:

- ***application of high pass filtering (300 Hz or above) and low pass filtering (below 3000 Hz)*** for obtaining low-frequency activity to observe spikes.
- ***application of single-threshold or double-threshold-based approaches*** to defining spike peaks. The spikes are selected based on the data points over a predefined threshold. In general, this threshold is indicated by the standard deviation of the recording. In our analysis, the data points exceed the 4th time the standard deviation of recording compares to baseline activity, the mean. The double threshold method is adopted for identifying both positive and negative spikes.
- ***extraction of features*** from detected spiking activity. Wavelet coefficients from each spike are calculated using a four-level multiresolution decomposition using a specific wavelet function. Several wavelet coefficients are obtained for isolated single-unit activity (SUA). Each wavelet coefficient here would characterize the spike shapes at different scales and times. Features need to be selected in a way helping the discrimination of the spiking activity for individual neurons.

- ***clustering single unit*** activities based on a similarity metric for differentiating the activities of different neurons. K-means clustering approach is used for grouping the spikes of different neurons with the adopted spike sorting algorithm method.

Spike detection and sorting are conducted using MATLAB ToolBox Wave_Clus[5]. At the end of the offline spike sorting process, 662 SUA (single unit activity) were detected from 1091 tracks in total. The medial trajectory has the lowest number of isolated neurons across depths in both hemispheres. The distributions of detected SUA per track are demonstrated in Table 4.

| | Depth | Left Hemisphere | | | | | Right Hemisphere | | | | | Total |
|---|---|---|---|---|---|---|---|---|---|---|---|---|
| | | A | C | L | M | P | A | C | L | M | P | |
| SINGLE DEPTH LEVEL | +3.5 | 0 | 3 | 0 | 0 | 0 | 0 | 0 | 0 | 0 | 0 | 3 |
| | +3 | 0 | 3 | 1 | 0 | 1 | 0 | 0 | 0 | 0 | 0 | 5 |
| | +2 | 2 | 2 | 7 | 0 | 1 | 2 | 1 | 0 | 1 | 0 | 16 |
| | +1 | 2 | 3 | 5 | 0 | 6 | 5 | 6 | 3 | 1 | 7 | 38 |
| | 0 | 6 | 1 | 4 | 0 | 5 | 8 | 9 | 2 | 3 | 4 | 42 |
| | -1 | 5 | 18 | 5 | 0 | 4 | 8 | 6 | 5 | 5 | 6 | 62 |
| | -2 | 10 | 4 | 6 | 2 | 3 | 10 | 9 | 3 | 1 | 8 | 51 |
| | -3 | 9 | 10 | 7 | 3 | 6 | 10 | 14 | 5 | 6 | 6 | 76 |
| | -4 | 7 | 4 | 5 | 3 | 3 | 8 | 11 | 7 | 3 | 5 | 56 |
| | -5 | 3 | 5 | 9 | 1 | 4 | 2 | 7 | 2 | 2 | 4 | 39 |
| | -6 | 6 | 7 | 10 | 4 | 7 | 8 | 8 | 4 | 9 | 2 | 65 |
| | -7 | 6 | 5 | 3 | 1 | 8 | 10 | 11 | 9 | 3 | 7 | 63 |
| | -8 | 6 | 3 | 2 | 5 | 1 | 7 | 7 | 3 | 2 | 5 | 46 |
| | -9 | 11 | 4 | 7 | 3 | 6 | 9 | 6 | 2 | 5 | 7 | 60 |
| | -10 | 6 | 6 | 2 | 1 | 4 | 4 | 3 | 1 | 4 | 9 | 40 |
| | Total | 79 | 78 | 73 | 23 | 59 | 91 | 98 | 46 | 45 | 70 | 662 |

**Table 4 - SUA Distribution per Tracks**
Isolated single-unit activities across depths of the electrode trajectories for all patients. The distribution of isolated neurons differentiates between hemispheres and trajectories.

#### 3.3. SUA, MUA & BUA Definitions

SUA, BUA, and MUA components in the raw MER recording represent different physiological phenomena near the electrode. From this view, exploiting the physiological meaning and the relationship between these components can provide immense intuition about the nearby neural activity around electrodes.

The single-unit activity represents the section of the raw data that belongs to the spiking of a single neuron. The [-0.5 +2.5]ms time interval around the peak values of all spikes was extracted from raw data and stored as the neural activity of a single neuron. In literature, single-unit activity denotes the separable single-unit spike train (SU-ST), not the non-separable multi-unit spike train (MU-ST)[6]. In our recordings, we observed up to 5 different SUA in a single MER signal. The MU-ST activities are formed by spikes from many neurons, with shapes that cannot be separated because of a low signal-to-noise ratio. The neurons contributing to the multiunit activity are relatively close to the electrode (for their spikes to be detected), but not close enough to enable the clustering of their shapes[7]. MUA holds both distinct spiking activities from one or more separable single neurons and spikes emerging from multiple units that cannot be separated by spike sorting[6]. Background activity reflects the summation of activities of more distant neurons in the raw MER recording. In a way, BUA represents smaller sub-noise level spikes generated by the surrounding neuronal population[8]. Background activity generally is constructed by subtracting

MUA (combination of SU-ST and MU-ST) from the raw local field potential (LFP) signal. Therefore BUA is the spike-free signal region. After the removal of MUA & SUA activity from the raw signal, several operations[6] are applied to the remaining time series to define exact background activity: (1) filtering signal with zero-phase LFP digital filter (prevent from phase distortions) around 500 Hz, (2) full-wave rectification of filtered signal and (3) removing DC offset.

##### 3.4. Stability Criterion for Sorted Neurons

The stability of detected neurons was investigated before proceeding with further analysis. This stability check of neurons is performed to avoid adding neurons to the dataset that do not have predicted neural activity or spike morphology. The adopted stability criterion[9] checks four different aspects of sorted neurons. **(a)** More than 90% of the total area of the amplitude histogram had to be above the detection threshold. We suggest the temporal region in the raw recording, which shows distinguishable changes compare to the baseline to indicate spiking activity. For each spike, all the spiking peaks will be compared with the defined double-threshold. If 90% of detected spikes peak is above this threshold level, the next criteria are checked. **(b)** The mean waveform of clustered activity of a neuron needs to have a typical action potential shape with a prominent peak. This step is completed with visual inspection during spike sorting. **(c)** The percentage of spikes occurring within 3 ms of each other had to be less than 1%. This criterion makes sure we do not involve spiking activity that occurs in the refractory period of the neuron. **(d)** The number of spikes detected had to be more than 20. For meaningful statistical conclusions, this threshold is defined to select the neurons. This stability criterion was applied to all the isolated neurons. Only the neurons which are passed the requirements of the stability criterion are accepted for the next steps.

#### 4. Candidate Neural Biomarker Generation for Dystonia Characterization

In our research activity, we aim to localize and differentiate the neural behaviour between different areas inside GPi for primary dystonia patients. So, a set of candidate neural biomarker definitions are important to explain different aspects of pathological functioning in GPi. In the following subsections, we will provide more information regarding these markers.

##### 4.1. Firing Rate and Regularity Related Neural Biomarkers

We defined 8 different candidate neural biomarker definitions which are related to the firing rate and firing regularity of neurons. The information regarding these candidate neural biomarkers is given in Table 5.

###### 4.1.1. Instantaneous and Mean Firing Rate

Instead of its usefulness, the mean firing rate does not provide a complete picture of the temporal firing characteristic of a neuron. Consequently, researchers generally rely on other types of schemes for a better understanding of neural work. An example of these schemes is the instantaneous firing rate of a neuron. In our study, we adopted Adaptive Kernel Smoother[10] (BAKS) to estimate the instantaneous firing rate. BAKS uses a kernel smoothing technique with an adaptive bandwidth. The neural process dynamics are encapsulated by the adaptive bandwidth for the selected kernel function. In this case, if the firing rate increases in the spike train, the bandwidth of the kernel function will be decreased by BASK. The adaptation of the bandwidth paves the way for defining accurate estimation of instantaneous firing rate under slow and fast-spiking dynamics. The kernel bandwidth is recognized by BAKS as a random variable with a prior distribution. It uses an empirical Bayesian method to change the posterior bandwidth. We set the $\alpha = 4$ and $\beta = N^{4/5}$ as two parameters of BAKS and estimated the instantaneous firing rate for each SUA.

#### 4.1.2. Inter Spike Interval

Since ISI provides information about the patterning of spiking activity, we defined four different neural biomarkers for indicating the aspects of inter-spike interval. The temporal distance between two subsequent spikes is called an inter-spike interval (ISI). ISI distribution of a spike train is constructed for a general understanding of temporal spike patterning. The different physiological phenomenon reflects themself in the ISI distribution of SUA, hence we want to use ISI distribution features as some of our candidate neural biomarkers. The distribution of the ISI is determined based on the bin size definition, and we set it as 10 ms. After ISI distribution generation we tried to fit each ISI distribution with gamma distribution which is a type of continuous probability distribution with two parameters: scale(λ) and shape(κ) parameters. For fitting the gamma distribution on ISI distribution, the native gamfit[11] function of MATLAB (Mathworks, Natick, MA, USA) was used. The proper fitting of the gamma distribution is essential for our study because the fitted shape and rate parameters are used for the definition of neural biomarkers. The Kolmogorov-Smirnov[12] (K-S Test) test was applied to the fitted distribution for measuring the goodness of fit. The fitted gamma distribution can be represented with two sets of parameters: shape and scale parameters. The following transformation is applied to get the shape and scale parameters of ISI distribution.

$$ISI_n = t_{n+1} - t_n$$

$$shape_{param} = e^{\kappa}$$

$$scale_{param} = \frac{1}{e^{\log(\lambda)+\kappa}}$$

| Candidate Neural Biomarker | Definition | Formulation |
|---|---|---|
| **firing_rate** | It is the simplest and most widely used neural biomarker for neural data analysis. It is directly connected to the outcome of neural functioning. | the rate parameter (λ) of the fitted gamma distribution for the ISI distribution |
| **regularity** | This metric quantifies the regularity of spiking of a neuron. This regularity metric provides intuition about the general firing characteristic of the neuron. Similar to the mean firing rate, the firing regularity is derived from the fitted parameters of ISI gamma distribution. Firing regularity[13] metrics also can help us to classify the neurons into subcategories based on their firing characteristic. In this study, the neurons are divided into three main categories: busting, tonic, and irregular. | $firing\ regularity = log(\kappa)$ <br> $firing\ pattern = \begin{cases} log(\kappa) \leq -0.3, & bursting \\ log(\kappa) \geq 0.3, & tonic \\ -0.3 > log(\kappa) > 0.3, & additional\ criterion \end{cases}$ <br> $additional\ criterion = \begin{cases} 70\%\ samples > \lvert \lambda \pm 0.5\lambda \rvert, bursting \\ 70\%\ samples < \lvert \lambda \pm 0.5\lambda \rvert, irregular \end{cases}$ |
| **cv** | The coefficient of variation in spike trains is evaluated to quantify the width of the ISI distribution[14]. It acts as a sort of measure for spike train irregularity. | $cv = \frac{\sqrt[2]{variance(ISI)}}{mean(ISI)}$ |
| **lv** | It is a metric originally defined to determine the intrinsic temporal dynamics of neuron spike trains. LV compares temporal variations with their local rates, and it is specifically defined for nonstationary processes. In the equation, each $\tau$ value indicates the timing of an observed spike where N parameters represent the total number of spikes in a spike train. Compared to the CV, it provides more robust results for distinguishing the activity of different neurons[15] | $lv = \frac{3}{N-2}\sum_{n=0}^{N-1}\left(\frac{(\tau_{n+1}-\tau_n)-(\tau_n-\tau_{n-1})}{(\tau_{n+1}-\tau_n)-(\tau_n-\tau_{n-1})}\right)^2$ |

| | | |
|---|---|---|
| **isi_mean** | The mean value of ISI represents the average temporal distance between two subsequent spikes of a neural structure. It is closely correlated with the mean firing rate biomarker. | $ISI_{mean} = shape_{param} \times scale_{param}$ |
| **isi_std** | To define the dispersion around the mean of ISI distribution, the standard deviation was calculated and used as a possible biomarker. This biomarker indicated the uncertainty of the duration between two subsequent spikes in the spike train. Intuitively, the higher dispersion around the ISI means indicates a wider range of temporal distance between spikes in spike train. | $ISI_{std} = \sqrt{shape_{param} \times scale_{param}{}^2}$ |
| **isi_skewness** | Skewness is a measure of the asymmetry of a distribution function and how the fitted gamma function deviates from a normal distribution. From a neural point of view, skewness represents how ISI values are symmetrically distributed around the mean of the distribution. | $ISI_{skewness} = \frac{2}{\sqrt{shape_{param}}}$ |
| **isi_rho** | Correlations between inter-spike intervals are occurred due to a combination of intrinsic mechanisms and the temporal properties of the input stimulus. ISI correlation can provide indirect information from bursting to periodical phase-locked firing[16]. ISI correlation is computed with the serial correlation coefficient formula. | $ISI_{rho} = SCC = \frac{\frac{1}{N}\sum_{i=1}^{N}(ISI_i - \overline{ISI_i}) \times (ISI_{i+k} - \overline{ISI_{i+k}})}{\frac{1}{N}\sum_{i=1}^{N}(ISI_i - \overline{ISI_i})^2}$ |

**Table 5 – Firing Rate and Firing Regularity-Related Neural Biomarker Definitions**
The names, definitions, and formulations of firing rate and firing regularity neural biomarkers.

#### 4.2. Neural Bursting

Bursting is a dynamic state where a neuron repeatedly fires discrete groups or bursts of spikes in a small temporal region. Bursting activity is thought to be involved in a variety of physiological processes, such as synapse formation, neural network connectivity, and long-term potentiation[17,18]. Bursts are also been considered as either physiological or pathological phenomena and potentially reflect the periodic oscillatory activity. In our case, dystonia-induced bursting activity is observed several studies. It was demonstrated that there was a subpopulation of theta-oscillatory cells with unique bursting characteristics inside GPi of cervical dystonia patients[19]. That is why we tried to characterize the bursting activity for dystonic GPi. There is a vast number of available burst detection in the existing literature. We used an approach called Rank Surprise[20] for the detection of bursting activity. In this method, the ISI values are calculated and ordered with ascending trends. The ordered ISI values are given a rank. The minimum ISI value takes the rank value equals 1.

$$ISI_n = t_{n+1} - t_n$$

$$R_n = rank(ISI_n) \; where \; min(rank(ISI_n)) = 1$$

Rn has a uniform distribution between 1 to N. That is why the algorithm is distribution-free because of giving a rank value to each ISI interval. Intuitively, a burst is a sequence of Rn values with successive low values. RS value calculated for checking a time interval burstiness. Rank Surprise values are calculated based on the log-likelihood of discrete uniform sum distribution P.

$$RS = -log(P(T_q < u))$$

$$where \; P(T_q < u) \; is \; discrete \; uniform \; sum \; distribution$$

If the RS value is higher than the user-defined threshold, the temporal region will be registered as busting activity. These regions will be subtracted from the spike train and the new iteration for finding the bursting region will be initiated until there is no bursting region detected by the algorithm. The bursting activity of all selected (the selection criteria for application of the Rank Surprise method shall be described in the firing regularity subsection) neurons. The number of detected bursts by the RS method is directly related to the distribution of surprise values. As authors of the paper suggested that some nonstationary in the ISI time distribution may dramatically affect the performance of the method. The mentioned recording merge criterion for stationarity, and variance coefficient, also makes this algorithm work more robust. The authors also suggested that the use of the RS method for multiunit recordings causes inaccurate results. As expected, near coincident spikes from different units will be resulted in very short ISI in the multiunit recording, whereas no burst is present. Two parameters are user-defined: the largest ISI allowed in a burst (limit) and the minimum RS (threshold) that a burst must have to be considered valid. We selected 75% as the largest ISI allowed in a burst and 0.01 as the minimum significance level selected burst region.

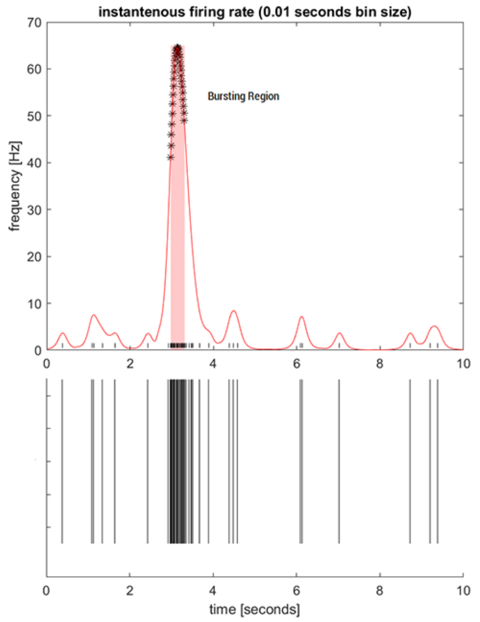


**Figure 2 - Bursting Activity Detected by Rank Surprise Method**
Two plots represent the neural activity. The first plot presents the firing rate of the neurons in the temporal domain and highlights the bursting activity of that neuron. A raster plot of the same neuron is illustrated in the second plot.

| Candidate Neural Biomarker | Definition | Formulation |
|---|---|---|
| **bspike_proportion** | It represents the proportion of spikes that are located inside the burst intervals compared to all existing spikes in the spike train. | $bspike\_proportion = \frac{\sum_{n=1}^{N_{bursts}} count_n(spikes)}{count(all\ spikes)}$ |
| **burst_avg_spikes** | The marker indicates the average number of spikes within the bursting interval. | $burst\_avg\_spikes = \frac{1}{N_{bursts}} \sum_{n=1}^{N_{bursts}} count_n(spikes)$ |
| **burst_index** | The burst index[21] is a metric that is defined for the assessment of possible changes in firing patterns. It is a widely used feature for cortical neural analysis. | $burst\ index = \frac{mean(ISI)}{mod(ISI)}$ |
| **interbi** | It indicates the average temporal distance between subsequent bursts. | $interbi = \frac{1}{N_{bursts} - 1} \sum_{n=1}^{N_{bursts}} (\tau_{burst\_start_{n+1}} - \tau_{burst\_finish_n})$ |

| | | |
|---|---|---|
| **intrabf** | The mean firing rate during the bursting periods within bursting intervals. | $intrabf = \frac{1}{N_{bursts}} \sum_{n=1}^{N_{bursts}} \frac{count_n(spikes)}{(\tau_{finish_n} - \tau_{start_n})}$ |
| **intrabi** | The mean interval of bursting activity within the spike train is set as a biomarker. | $intrabi = \frac{1}{N_{bursts}} \sum_{n=1}^{N_{bursts}} (\tau_{burst\_finish_n} - \tau_{burst\_start_n})$ |

**Table 6 – Neural Bursting Related Neural Biomarker Definitions**
The names, definitions, and formulations of bursting-related neural biomarkers.

#### 4.3. Power Spectrum

Power spectrum components of single-unit activity are also considered for the candidate neural biomarker generation process. Firstly, we extracted the action potentials of single-unit activity from raw MER recording and remove the mean to get rid of the power at f=0. In the following step, we removed the powerline noise (50 Hz) and its first five harmonics to prevent introducing bias to our power spectrum and neural oscillation estimation. We adopted the Welch PSD estimation by selecting the Hanning window as our tapering function with a window size equal to the data length divided by 20. The overlapping of the windows is selected as 0.5. In the last step of power spectrum estimation, we normalize the power spectrum by dividing the power values in each frequency by the total power of the SUA. This normalization procedure is essential for acquiring a comparable power spectrum across all isolated single units. It is a common approach to studying neural oscillations in delta (1-4 Hz), theta (4-8 Hz), alpha (8-12 Hz), beta (12–30 Hz), and gamma (30-100 Hz) frequency bands. Hence, we generated three individual neural biomarkers per frequency band:

- the minimum power in the frequency band
- the mean power of the frequency band
- the maximum power in the frequency band

#### 4.4. Neural Oscillations

Neural oscillations are rhythmic patterns of neural activity in the CNS. The generation of neural oscillations can be mediated by multiple factors such as intrinsic properties of the oscillating neurons or network-wide interactions between other neurons. From this perspective, this rhythmic activity of neurons plays a major role in neural processing and behavior[22]. Instead of the normal functioning of neurons, neural oscillations can also appear in pathological conditions. Pathological synchronous activity within the corticothalamic-basal ganglia network has been identified as a key hallmark of Parkinson's disease[23]. The standard approach for defining the oscillatory neurons is achieved by investigating the power spectral density (PSD) of SUA. The neural oscillations manifest themselves with prominent peaks in their PSD. For each of these frequency bands, the following neural biomarkers were evaluated:

- peak power in the frequency band
- location of the peak in terms of corresponding frequency

These 10 neural biomarkers are believed to reflect the complete view of the oscillatory characteristic of GPi neurons for primary dystonia patients on a microscopic scale. Additionally, we adopt a criterion that evaluates the strength of the oscillation. The baseline magnitude is selected as the median magnitude of the whole spectrum. The confidence of each peak in the frequency band of interest is defined based on the threshold value. If the peak magnitude of the frequency band is higher than the three interquartile ranges compared to the baseline magnitude, the oscillation in that frequency band is considered significant and we involved only significant oscillations in our analysis.

#### 4.5. Spectral Coherence Between BUA & SUA

The coupling between time series can be investigated with various methods, either in frequency or time domain. Spectral coherence is a commonly used method for exploiting functional connectivity between different brain regions as a function of frequency. The oscillation phase of the network has a significant impact on the spiking probability of neurons within a neural population[24]. As a result, the frequency of oscillation cycles is proportional to the ease of excitability. In neuroscience, estimating spectral coherence is frequently used to measure phase-locking strength between two neuronal populations[25]. SUA and BUA represent two distinct neural populations around MER electrodes (Figure 3). From this perspective, investigating the phase-locking strength between these two neuron populations across all tracks can provide inside into the pathological functioning of dystonic GPi. Coherence is the normalized version of the cross power spectrum density of two time series. Spectral coherence has multiple formulations. In the literature, the quadratic / magnitude squared coherence is widely used. Based on the definition, the square of cross power spectral density is divided by the multiplication of the marginal power spectral densities.

$$Coh_{BUA,SUA}\,(f) = \frac{|P_{BUA,SUA}(f)|^2}{P_{BUA,BUA}(f)P_{SUA,SUA}(f)}$$

An important aspect related to spectral coherence is the significance of calculated coherence for each frequency. Since the spectral coherence does not follow any distribution, there is no comparison of the coherence for testing the hypothesis. The solutions are provided for this issue; conducting surrogate data analysis to generate a reference distribution or using one of the theoretical approximation formulations for measuring the significance in the neuroscience literature. Due to the lower computation time and cost, the second option is chosen.

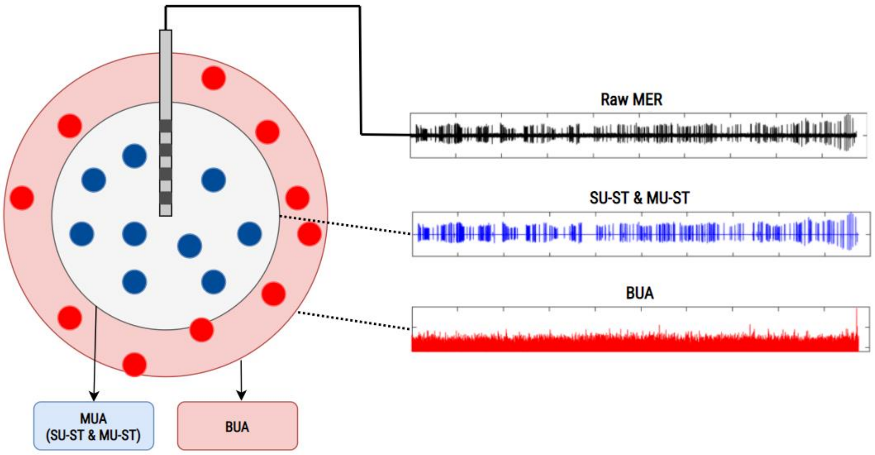


**Figure 3 - Coherence Schema for SUA & BUA**
A schema demonstrates the active stimulation sites of the DBS electrode and what kind of signal components are extracted from different stimulation sites.

In the literature, the following formulation is found to define the significance of calculated spectral coherence. The α (alpha) value represents the adopted significance value which we defined as 0.95. During the spectrum estimation by the Welch method, the PSD is estimated by defining a windowing function with a specific length. L parameter represents the number of windowing functions without non-overlapping.

$$Coherence_{limit} = 1 - (1 - \alpha)^{\frac{1}{L-1}}$$

Multiple coherence-related neural biomarkers were defined for 5 different frequency bands. The native MATLAB functions were used to detect peaks in each band. Based on these calculations, four different candidate neural biomarkers are defined for each of these bands:

- the mean coherence of the frequency band,
- the mean of only significant coherence of the frequency band
- the value of peak coherence value within the frequency band,
- the frequency that has the peak coherence inside the selected frequency band.

For investigating the functional structure of the dystonic GPi, we defined 54 distinct candidate neural biomarkers.

#### 5. Mutual Information Analysis

The entropy of a random variable is a metric that quantifies a random variable's unpredictability. The scope of entropy can be extended to mutual information. Mutual information is a non-linear metric that tries to quantify the coupling between two variables. It evaluates how much information is transmitted about another variable on average in the selected variable. Briefly, it indicates how much one random variable can inform you about another random variable. If the two variables are independent, seeing the outcome of one has no bearing on the uncertainty of the other, and therefore the mutual information between them is zero. From this perspective, investigating the values of neural biomarkers with different spatial variables will help us to determine the non-linear relationship between them. In this work, mutual information was investigated in two different domains: 3D MNI space and 1D relative depths of microelectrode recordings.

#### 5.1. Performance Metrics

One of the most important aspects of the success of machine learning applications is the selection of the right performance metric. In our study, we have two types of classification procedures: binary and multiclass classification. Both the two classifications: GPi in-out and GPi dorsal-ventral in MNI space and GPi in-out classification in relative depths are examples of binary classification. For these binary classifications, we have selected the following performance metrics: balanced accuracy, weighted F1, and weighted AUC score.

$$precision = \frac{True_{positives}}{True_{positives} + False_{positives}}$$

$$sensitivity = recall = \frac{True_{positives}}{True_{positives} + False_{negatives}}$$

$$specificity = \frac{True_{Negatives}}{True_{Negatives} + False_{positives}}$$

$$balanced_{accuracy} = \frac{sensitivity + specificity}{2}$$

$$F1 = 2\,x\,\frac{precision\ x\ recall}{precision\ x\ recall}$$

Accuracy is the simplest performance metric out of all metrics which can provide the wrong intuition about the performance of the classifier, especially in imbalanced class problems. Hence, we used the "balanced" version of the accuracy metric. F1 is a widely popular metric for binary classification because of combining two other metrics (precision and recall) to get a composite evaluation metric. Since we are giving the same importance to each class across all classification tasks, instead of using the F1 score, we used the weighted-F1 score where we calculated the F1 score for each class and then took the weighted average of these F1 scores based on the support of the class. The degree of separability of classes is represented by the AUC score. In another way, the AUC indicates how well the model predicts negative classes like negative and positive courses as positive. To get the AUC score, the TPR and FPR are needed to be defined. Similar to the weighted F1 score, we also adopted the weighted version of the AUC score.

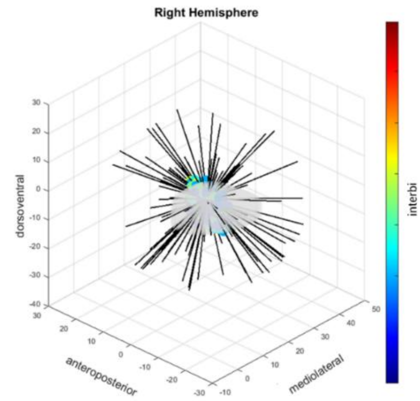


**Figure 4 – Directions with Significant Mutual Information for Mean Inter-Burst Interval Biomarker**
The isotropic behaviour of mean inter-burst interval biomarker in terms of significant mutual information calculated for dorsoventral localization task in 3D MNI reference frame.

### 6. Feature Selection Result

#### 6.1. Defining Cut-Off Points for Feature Selection Steps

For the first step of feature selection, we computed the mutual information and conducted the bootstrap test with 500 iterations to draw our sampling distribution. Assuming the normality of the bootstrap distribution, we defined the significance value as z-score≥2 (p≤0.05) for the mutual information of each neural biomarker to accept them for the second step of the feature selection.

In the second step, we used the set of biomarkers that were selected in the first step. Subsequently, we proceed with the following substeps to define our threshold and significant biomarker set for the localization task:

- SS1: Compute the information gained from each neural biomarker by constructing a Random Forest Tree.
- SS2: Compute the decoding performance in the prediction block tasks considering a set including all biomarkers
- SS3: Check if the performances of the biomarker set are robust in the prediction block (If there is lateralization; the performance agreement between right and left hemisphere tasks with a lower performance in merged hemisphere task is expected. If there is no lateralization, we expected similar performance across all three tasks in the prediction block.) check the robustness in the tasks of prediction block (If there is lateralization, cross-validation tasks are expected to have low performance and agreement in leave-one-trajectory-out tasks. If there is no lateralization, we expected similar performance across all seven tasks in the prediction block).
- SS4: repeat SS2-SS3 restricted to the subset of biomarkers including only those with information gain above a cut-off value TH from the heuristically selected set [0.005, 0.010, 0.015, 0.020, 0.025, 0.030, 0.035, 0.040, 0.045, 0.050, 0.055,0.060, 0.065, 0.070].
- SS5: Find the subset of biomarkers leading to optimal robust performance. The associated TH is the information gain cut-off.

**Feature Selection Step 2: Information Gain Threshold Selection Procedure**

**input** $\boldsymbol{reference_{frame}} \in [1D, 3D]$
$\boldsymbol{classification} \in [GPi_{in-out}, GPi_{dorsoventral}]$
$\boldsymbol{task_{validation\_block}} \in [hemisphere_{merged}, hemisphere_{right}, hemisphere_{left}]$
$\boldsymbol{task_{prediction\_block}} \in [cross\_hemisphere_{right}, cross\_hemisphere_{left},$
$leave\_trajectory\_out_{anterior}, leave\_trajectory\_out_{central}, leave_{trajectory_{out_{lateral}}},$
$leave\_trajectory\_out_{medial}, leave\_trajectory\_out_{posterior}]$
$array\ \boldsymbol{biomarkers}\ of\ \boldsymbol{n}\ string$
$array\ \boldsymbol{biomarkers_{IG}}\ of\ \boldsymbol{n}\ doubles$

$threshold_{set} \leftarrow [0.01, 0.015, 0.02, 0.025, 0.03, 0.035, 0.04, 0.045, 0.05, 0.06, 0.065, 0.07]$
$cut_{off} \leftarrow \mathrm{threshold_{set}}[1]$
$auc_{optimal} \leftarrow 0$
*significant_biomarkers←[ ]*

**for** *i←1* **to** $length(threshold_{set})$ **do**
  $threshold \leftarrow threshold_{set}[i]$
  *biomarker_list←[ ]*
  **for** *j←1* **to** *n* **do**
    $biomarker_{IG} \leftarrow biomarkers_{IG}\ [i]$
    **if** $biomarker_{IG} \geq threshold$ **then**
      *biomarker_list[end]←biomarkers[j]*

  $auc_{merged}$ ←***localizationPerformance***$(classification,\ hemisphere_{merged},\ biomarker\_list)$
  $auc_{right}$ ←***localizationPerformance***$(classification, hemisphere_{right},\ biomarker\_list)$
  $auc_{left}$ ←***localizationPerformance***$(classification, hemisphere_{left},\ biomarker\_list)$
  $robust_{validation\_block}, lateralization = \boldsymbol{validationBlockRobustness}(auc_{merged}, auc_{right}, auc_{left})$

  **if** $robust_{validation\_block} = true$ **then**
    **if** $lateralization = true$ **then**
      $robust_{prediction\_block} \leftarrow \boldsymbol{predictionBlockRobustness}\ (classification,\ task_{prediction\_block},$
      $biomarker_{list},\ auc_{right})$
    **else then**
      $robust_{prediction\_block} \leftarrow \boldsymbol{predictionBlockRobustness}\ (classification,\ task_{prediction\_block},$

$biomarker\_list,\ auc_{merged})$
**if** $robust_{prediction\_block} = true$ **then**
  **if** $lateralization = true$ **then**
    **if** $auc_{right} \geq auc_{optimal}$ **then**
      $auc_{optimal} = auc_{right}$
      $cut_{off} \leftarrow threshold$
      $significant\_biomarkers \leftarrow biomarker\_list$
  **else then**
    **if** $auc_{merged} \geq auc_{optimal}$ **then**
      $auc_{optimal} = auc_{merged}$
      $cut_{off} \leftarrow threshold$
      $significant\_biomarkers \leftarrow biomarker\_list$

Due to the complex nature of this subsection of the pipeline, here we present the pseudocode of threshold defining procedure for the second step of feature selection. Since the cut-off definition is derived from the performance of a set of features, and not single features, there was no need to apply multiple comparison corrections. Hence, the definition of our cross-validation tasks helped us to determine the optimum set of neural biomarkers and cut-off values for each localization. For GPi in-out localization in the 1D and 3D reference frame and GPi dorsoventral localization in the 3D reference frame, we found the optımal cut-off value to be 0.05. The optimal threshold point for GPi in-out localization in the 3D reference frame we found the optımal cut-off value to be 0.03.

#### 6.2. Neural Biomarkers for GPi in-out Localization in 1D

| Feature | Feature Selection 1st Step | | | | Feature Selection 2nd Step |
|---|---|---|---|---|---|
| | Mutual Information | Z-Score | Bootstrap Mean | Bootstrap Standard Deviation | Mean Decrease in Total Entropy |
| firing_rate | 0.03924 | 3.53923 | 0.00311 | 0.01021 | 0.042093 |
| cv | 0.03383 | 2.92200 | 0.00362 | 0.01034 | 0.062429 |
| isi_mean | 0.03967 | 3.49479 | 0.00345 | 0.01036 | 0.042329 |
| isi_std | 0.04272 | 3.87863 | 0.00393 | 0.01000 | 0.057545 |
| İntrabf | 0.03354 | 2.61559 | 0.00682 | 0.01022 | 0.017214 |
| İnterbi | 0.04765 | 4.00772 | 0.00675 | 0.01021 | 0.015370 |
| burst_avg_spikes | 0.03178 | 2.22446 | 0.00835 | 0.01053 | 0.016160 |
| delta_band_mean_power | 0.05476 | 5.03064 | 0.00306 | 0.01028 | 0.041697 |
| delta_band_max_power | 0.04259 | 3.58223 | 0.00359 | 0.01089 | 0.050061 |
| delta_band_min_power | 0.04786 | 4.26317 | 0.00363 | 0.01038 | 0.050359 |
| theta_band_mean_power | 0.04478 | 3.90063 | 0.00414 | 0.01042 | 0.041190 |
| theta_band_max_power | 0.05748 | 5.22345 | 0.00380 | 0.01028 | 0.043302 |
| theta_band_min_power | 0.05551 | 5.01112 | 0.00377 | 0.01032 | 0.051437 |
| alpha_band_mean_power | 0.05406 | 4.93120 | 0.00376 | 0.01020 | 0.037910 |
| alpha_band_max_power | 0.04472 | 3.90169 | 0.00358 | 0.01055 | 0.038359 |
| alpha_band_min_power | 0.07433 | 6.73970 | 0.00365 | 0.01049 | 0.050350 |
| beta_band_mean_power | 0.06395 | 5.95098 | 0.00380 | 0.01011 | 0.042108 |
| beta_band_max_power | 0.04703 | 4.27481 | 0.00316 | 0.01026 | 0.042381 |
| beta_band_min_power | 0.05765 | 5.17704 | 0.00332 | 0.01049 | 0.042063 |
| gamma_band_mean_power | 0.04945 | 4.42030 | 0.00373 | 0.01034 | 0.043737 |
| gamma_band_max_power | 0.03778 | 3.25407 | 0.00370 | 0.01047 | 0.043269 |
| gamma_band_min_power | 0.05880 | 5.03240 | 0.00354 | 0.01098 | 0.051960 |

| | | | | | |
|---|---|---|---|---|---|
| delta_band_peak_coherence | 0.02870 | 2.34981 | 0.00439 | 0.01035 | 0.022294 |
| delta_band_peak_coherence_frequency | 0.02362 | 1.78176 | 0.00523 | 0.01032 | 0.027913 |
| theta_band_mean_significant_coherence | 0.02613 | 1.94223 | 0.00608 | 0.01032 | 0.026470 |

**Table 7. Two-step neural biomarker selection results of GPi in-out neuron localization for merged hemispheres task in the 1D relative depth reference frame.** The table presents the results of the feature selection process of GPi in-out localization in the 1D reference frame. Only features that show significant mutual information in the first step of feature selection are presented in this table. Green highlighted rows indicate biomarkers selected by the process for this task.

#### 6.3. Information Gain of Neural Biomarkers for GPi in-out Localization in 3D

| feature | Feature Selection 2nd Step | |
|---|---|---|
| | Mean Decrease in Total Entropy (Left Hemisphere) | Mean Decrease in Total Entropy (Right Hemisphere) |
| firing_rate | 0.018276 | 0.019836 |
| regularity | 0.024069 | 0.035302 |
| isi_rho | 0.041931 | 0.024269 |
| cv | 0.026351 | 0.029147 |
| lv | 0.020962 | 0.030842 |
| isi_mean | 0.018815 | 0.027377 |
| isi_std | 0.020622 | 0.026174 |
| isi_skewness | 0.028430 | 0.035220 |
| intrabf | 0.008383 | 0.005407 |
| interbi | 0.008283 | 0.005364 |
| intrabi | 0.005671 | 0.004545 |
| burst_index | 0.005792 | 0.004501 |
| burst_avg_spikes | 0.005315 | 0.005663 |
| bspike_proportion | 0.030388 | 0.005339 |
| delta_band_mean_power | 0.025846 | 0.015725 |
| delta_band_max_power | 0.025421 | 0.016664 |
| delta_band_min_power | 0.016707 | 0.015601 |
| theta_band_mean_power | 0.016662 | 0.017493 |
| theta_band_max_power | 0.022713 | 0.020280 |
| theta_band_min_power | 0.020003 | 0.019015 |
| alpha_band_mean_power | 0.025728 | 0.019791 |
| alpha_band_max_power | 0.033527 | 0.015391 |
| alpha_band_min_power | 0.015855 | 0.013681 |
| beta_band_mean_power | 0.018312 | 0.014372 |
| beta_band_max_power | 0.024497 | 0.021982 |
| beta_band_min_power | 0.017802 | 0.011504 |
| gamma_band_mean_power | 0.023969 | 0.018142 |
| gamma_band_max_power | 0.025478 | 0.022859 |
| gamma_band_min_power | 0.021654 | 0.024505 |
| oscillation_gamma_exist | 0.003129 | 0.001899 |
| oscillation_gamma_freq | 0.014296 | 0.007317 |
| oscillation_gamma_power | 0.006531 | 0.005034 |
| delta_band_mean_coherence | 0.023263 | 0.039634 |
| delta_band_mean_significant_coherence | 0.010289 | 0.014204 |
| delta_band_peak_coherence | 0.005481 | 0.028225 |
| delta_band_peak_coherence_frequency | 0.007064 | 0.014217 |
| theta_band_mean_coherence | 0.022701 | 0.036155 |

| | | |
|---|---|---|
| theta_band_mean_significant_coherence | 0.003403 | 0.008569 |
| theta_band_peak_coherence | 0.022572 | 0.017053 |
| theta_band_peak_coherence_frequency | 0.012750 | 0.012727 |
| alpha_band_mean_coherence | 0.026219 | 0.030651 |
| alpha_band_mean_significant_coherence | 0.009309 | 0.008245 |
| alpha_band_peak_coherence | 0.009078 | 0.025328 |
| alpha_band_peak_coherence_frequency | 0.015217 | 0.010938 |
| beta_band_mean_coherence | 0.029478 | 0.028396 |
| beta_band_mean_significant_coherence | 0.008561 | 0.032471 |
| beta_band_peak_coherence | 0.020195 | 0.026298 |
| beta_band_peak_coherence_frequency | 0.017246 | 0.029106 |
| gamma_band_mean_coherence | 0.042760 | 0.021745 |
| gamma_band_mean_significant_coherence | 0.029618 | 0.027162 |
| gamma_band_peak_coherence | 0.035536 | 0.025677 |
| gamma_band_peak_coherence_frequency | 0.027842 | 0.022957 |

**Table 8 – The neural biomarker selection results of GPi in-out localization of dystonic GPi neurons for separate hemispheres tasks of prediction block in the 3D MNI reference frame**. The table contains information regarding the results of entropy-based marker selection with 0.03 entropy criteria for GPi in-out localization task in 3D MNI coordinates.

#### 6.4. Neural Biomarkers for Dorsoventral Localization in 3D

| feature | 1st Step | | | | | | 2nd Step |
|---|---|---|---|---|---|---|---|
| | Preferential Direction | | | | | Significant Dorsoventral Direction Percentage | Mean Decrease in Total Entropy |
| | I | Z-Score | Bootstrap Mean | Bootstrap Standard Deviation | Significance | | |
| alpha_band_max_power | 0.08055 | 3.18442 | 0.00972 | 0.02224 | True | 24.3243 | 0.034125 |
| alpha_band_mean_coherence | 0.07473 | 2.76330 | 0.01648 | 0.02108 | True | 4.5045 | x |
| alpha_band_mean_power | 0.10937 | 4.56206 | 0.01381 | 0.02095 | True | 48.6486 | 0.034228 |
| alpha_band_mean_significant_coherence | 0.04820 | 1.15995 | 0.02331 | 0.02146 | False | 0 | x |
| alpha_band_min_power | 0.10977 | 4.60676 | 0.00926 | 0.02182 | True | 49.5495 | 0.037848 |
| alpha_band_peak_coherence | 0.05180 | 1.72319 | 0.01781 | 0.01973 | False | 0 | x |
| alpha_band_peak_coherence_frequency | 0.05385 | 1.81624 | 0.01578 | 0.02096 | False | 0 | x |
| beta_band_max_power | 0.07391 | 2.86983 | 0.01014 | 0.02222 | True | 9.9099 | x |
| beta_band_mean_coherence | 0.05544 | 1.97631 | 0.01140 | 0.02229 | True | 3.6036 | x |
| beta_band_mean_power | 0.10404 | 4.22344 | 0.01399 | 0.02132 | True | 31.5315 | 0.033894 |
| beta_band_mean_significant_coherence | 0.05624 | 1.78061 | 0.01474 | 0.02331 | True | 2.7027 | x |
| beta_band_min_power | 0.10725 | 4.34196 | 0.00880 | 0.02267 | True | 50.4505 | 0.039292 |
| beta_band_peak_coherence | 0.07546 | 3.04499 | 0.00829 | 0.02206 | True | 9.009 | x |
| beta_band_peak_coherence_frequency | 0.06281 | 2.37398 | 0.01283 | 0.02106 | True | 6.3063 | x |
| bspike_proportion | 0.07148 | 2.25402 | 0.02374 | 0.02118 | True | 3.6036 | x |
| burst_avg_spikes | 0.11535 | 4.47433 | 0.02526 | 0.02014 | True | 36.036 | 0.012435 |
| burst_index | 0.10407 | 3.80775 | 0.02464 | 0.02086 | True | 18.018 | 0.016578 |
| cv | 0.07555 | 3.00620 | 0.00866 | 0.02225 | True | 14.4144 | 0.066889 |
| delta_band_max_power | 0.10525 | 4.41486 | 0.00848 | 0.02192 | True | 26.1261 | 0.039661 |
| delta_band_mean_coherence | 0.06323 | 2.44735 | 0.01209 | 0.02090 | True | 1.8018 | x |
| delta_band_mean_power | 0.08875 | 3.58076 | 0.00965 | 0.02209 | True | 10.8108 | 0.035386 |
| delta_band_mean_significant_coherence | 0.06800 | 2.09451 | 0.02190 | 0.02201 | True | 1.8018 | x |
| delta_band_min_power | 0.10938 | 4.70894 | 0.01596 | 0.01984 | True | 34.2342 | 0.037654 |
| delta_band_peak_coherence | 0.09006 | 3.45918 | 0.01662 | 0.02123 | True | 24.3243 | 0.050941 |

| | | | | | | | |
|---|---|---|---|---|---|---|---|
| delta_band_peak_coherence_frequency | 0.09218 | 3.23446 | 0.01927 | 0.02254 | True | 7.2072 | x |
| firing_rate | 0.08405 | 3.46534 | 0.00898 | 0.02167 | True | 25.2252 | 0.058975 |
| gamma_band_max_power | 0.07378 | 2.78445 | 0.01659 | 0.02054 | True | 9.9099 | x |
| gamma_band_mean_coherence | 0.06668 | 2.39355 | 0.01578 | 0.02127 | True | 9.9099 | x |
| gamma_band_mean_power | 0.09176 | 3.56752 | 0.01457 | 0.02164 | True | 23.4234 | 0.030436 |
| gamma_band_mean_significant_coherence | 0.09098 | 3.42329 | 0.01750 | 0.02146 | True | 22.5225 | 0.042503 |
| gamma_band_min_power | 0.12005 | 4.94078 | 0.01206 | 0.02186 | True | 66.6667 | 0.047774 |
| gamma_band_peak_coherence | 0.08104 | 3.09067 | 0.01766 | 0.02051 | True | 15.3153 | 0.044908 |
| gamma_band_peak_coherence_frequency | 0.05876 | 2.33436 | 0.00795 | 0.02176 | True | 3.6036 | x |
| interbi | 0.08112 | 2.67053 | 0.02493 | 0.02104 | True | 18.9189 | 0.017362 |
| intrabf | 0.08441 | 2.82739 | 0.02498 | 0.02102 | True | 3.6036 | x |
| intrabi | 0.11995 | 4.38416 | 0.02535 | 0.02158 | True | 25.2252 | 0.013591 |
| isi_mean | 0.07618 | 2.97450 | 0.00941 | 0.02245 | True | 28.8288 | 0.050866 |
| isi_rho | 0.07637 | 3.03889 | 0.01350 | 0.02069 | True | 5.4054 | x |
| isi_skewness | 0.07315 | 2.61789 | 0.01712 | 0.02140 | True | 5.4054 | x |
| isi_std | 0.09094 | 3.68031 | 0.00967 | 0.02208 | True | 20.7207 | 0.062220 |
| lv | 0.07990 | 3.09426 | 0.01600 | 0.02065 | True | 9.009 | x |
| oscillation_beta_frequency | 0.04924 | 2.25866 | 0.02405 | 0.01115 | True | 1.8018 | x |
| oscillation_beta_power | 0.05329 | 2.96147 | 0.02389 | 0.00993 | True | 1.8018 | x |
| oscillation_delta_frequency | 0.03406 | 1.77424 | 0.02148 | 0.00709 | False | 0 | x |
| oscillation_delta_power | 0.03623 | 2.10891 | 0.02121 | 0.00712 | True | 0.9009 | x |
| oscillation_gamma_frequency | 0.05730 | 1.51460 | 0.02691 | 0.02007 | False | 0 | x |
| oscillation_gamma_power | 0.07652 | 2.60792 | 0.02602 | 0.01936 | True | 2.7027 | x |
| regularity | 0.07084 | 2.61990 | 0.01723 | 0.02046 | True | 4.5045 | x |
| theta_band_max_power | 0.10843 | 4.41451 | 0.00991 | 0.02232 | True | 37.8378 | 0.037619 |
| theta_band_mean_coherence | 0.06633 | 2.46973 | 0.01196 | 0.02201 | True | 9.9099 | x |
| theta_band_mean_power | 0.09925 | 4.27680 | 0.01086 | 0.02067 | True | 42.3423 | 0.036428 |
| theta_band_mean_significant_coherence | 0.06892 | 2.12929 | 0.02141 | 0.02231 | True | 0.9009 | x |
| theta_band_min_power | 0.08547 | 3.39133 | 0.01589 | 0.02052 | True | 43.2432 | 0.036732 |
| theta_band_peak_coherence | 0.12611 | 4.96943 | 0.01002 | 0.02336 | True | 11.7117 | 0.041225 |
| theta_band_peak_coherence_frequency | 0.08735 | 3.41939 | 0.01074 | 0.02240 | True | 16.2162 | 0.040430 |

**Table 9- Two-step neural biomarker selection results of dorsoventral localization for dystonic GPi neurons for merged hemispheres task in the 3D MNI reference frame.** The table contains information regarding preferential directions of candidate neural biomarkers and the results of each step of the feature selection process for the dorsoventral localization task.

### 7. Neuron Localization Results

#### 7.1. GPi In-Out Localization in 1D Relative Space

| Biomarker | Merged Hemispheres | | | Right Hemisphere | | | Left Hemisphere | | |
|---|---|---|---|---|---|---|---|---|---|
| | Cramér–Von Mises Test | | Mann-Whitney U Test (corrected) | Cramér–Von Mises Test | | Mann-Whitney U Test (corrected) | Cramér–Von Mises Test | | Mann-Whitney U Test (corrected) |
| | GPi-in | GPi-out | | GPi-in | GPi-out | | GPi-in | GPi-out | |
| **alpha_band_min_power** | 4.642 e-09 | 4.612 e-09 | 1 | x | x | x | x | x | x |
| **beta_band_max_power** | x | x | x | 3.981 e-09 | 2.863 e-10 | 0.922 | x | x | x |
| **cv** | 1.262 e-10 | 3.468 e-10 | 1 | 6.801 e-08 | 6.814 e-07 | 0.939 | 2.111 e-07 | 4.686 e-07 | 0.671 |
| **delta_band_max_power** | 4.630 | 4.610 | 1 | 3.9629 | 2.864 | 0.939 | x | x | x |

| | | | | | | | | | |
|---|---|---|---|---|---|---|---|---|---|
| | e-09 | e-09 | | e-09 | e-10 | | | | |
| **delta_band_min_power** | 4.612 e-09 | 4.642 e-09 | 1 | x | x | X | x | x | x |
| **delta_band_peak_coherence** | x | x | x | 3.865 e-09 | 2.099 e-09 | 0.006 | x | x | x |
| **delta_band_peak_coherence_frequency** | x | x | x | 7.065 e-11 | 7.281 e-09 | 0.039 | x | x | x |
| **firing_rate** | x | x | x | 1.821 e-09 | 6.225 e-10 | 0.922 | 2.528 e-09 | 2.256 e-09 | 0.0386 |
| **gamma_band_min_power** | 4.613 e-09 | 4.645 e-09 | 1 | x | x | X | x | x | x |
| **isi_mean** | x | x | x | 2.017 e-09 | 6.463 e-10 | 0.922 | 1.578 e-09 | 6.374 e-11 | 0.0386 |
| **isi_std** | 1.383 e-09 | 1.395 e-09 | 0.0425 | x | x | X | 7.825 e-10 | 2.272 e-11 | 0.0386 |
| **theta_band_min_power** | 4.612 e-09 | 4.643 e-09 | 1 | 3.983 e-09 | 2.866 e-10 | 0.939 | x | x | x |

**Table 10 – Results of Cramér–Von Mises and Mann-Whitney U Tests Applied on Selected Features for GPi In-Out Localization in 1D Relative Space (p-values are corrected according to Holm-Bonferroni correction)**

| Hemisphere | Performance Metric | Decision Tree Classifier | Random Forest Tree Classifier | K-Nearest Neighbors Classifier | Gaussian Process Classifier | Support Vector Machine Classifier | Voting Classifier (KNN+RF) |
|---|---|---|---|---|---|---|---|
| **merged** | balanced accuracy | 0.537 ±0.019 | 0.588 ±0.027 | 0.623 ±0.033 | 0.534 ±0.034 | 0.566 ±0.050 | 0.617 ±0.031 |
| | weighted-AUC | 0.575 ±0.026 | 0.619 ±0.051 | 0.692 ±0.043 | 0.580 ±0.053 | 0.595 ±0.051 | 0.691 ±0.044 |
| | weighted-F1 | 0.539 ±0.020 | 0.589 ±0.027 | 0.624 ±0.032 | 0.536 ±0.034 | 0.565 ±0.049 | 0.618 ±0.032 |
| **right** | balanced accuracy | 0.590 ±0.065 | 0.598 ±0.038 | 0.638 ±0.039 | 0.607 ±0.053 | 0.571 ±0.030 | 0.633 ±0.048 |
| | weighted-AUC | 0.627 ±0.041 | 0.652 ±0.056 | 0.708 ±0.030 | 0.656 ±0.034 | 0.619 ±0.018 | 0.705 ±0.039 |
| | weighted-F1 | 0.588 ±0.066 | 0.595 ±0.038 | 0.635 ±0.042 | 0.608 ±0.051 | 0.568 ±0.027 | 0.630 ±0.048 |
| **left** | balanced accuracy | 0.572 ±0.042 | 0.597 ±0.076 | 0.608 ±0.039 | 0.536 ±0.056 | 0.562 ±0.057 | 0.636 ±0.048 |
| | weighted-AUC | 0.578 ±0.045 | 0.619 ±0.036 | 0.681 ±0.016 | 0.578 ±0.058 | 0.564 ±0.047 | 0.664 ±0.022 |
| | weighted-F1 | 0.569 ±0.039 | 0.597 ±0.078 | 0.608 ±0.039 | 0.460 ±0.113 | 0.542 ±0.060 | 0.637 ±0.048 |

**Table 11 – Performances of Trained Classifiers for GPi In-Out Localization in Main Analysis Modes**

The individual performances of the Decision Tree, Random Forest Tree, KNN, Gaussian Process, and SVM classifiers for GPi in-out localization along with the Voting classifier that was constructed based on soft-voting using the most successful two individual classifiers in the relative depth domain.

The results of GPi in-out localization in the relative depth domain state that the KNN is the most successful algorithm across all three main analysis modes. The Random Forest algorithm shows the second-best performances for merged and left hemisphere analyses. Even though Gaussian Process Classifier has better performance than that of the Random Forest Trees model in the right hemisphere analysis, it generally showed the weakest performance in the remaining analyses. As a result, we selected trained KNN and Random Forest Tree classifiers to construct Voting Classifiers with a soft voting mechanism. Nonetheless, the performances of KNN and Voting classifiers are quite similar but the robustness of the Voting Classifier is clearer in the cross-validation tasks. The results of seven cross-validation analyses in this task are given in Table 9. Cross-hemispheric cross-validation tasks are conducted with a hemisphere-only trained model. For example, the model of right hemisphere GPi is trained with 350 neurons detected in the right hemisphere. This trained model is tested with the remaining 312 left hemisphere neurons and the performance metrics are calculated. In terms of the leave-one-out cross-validation task, we used the model of merged hemisphere main analysis mode. We trained the model with the neurons of four trajectories. The remaining trajectory is saved for the testing procedure.

| Performance Metric | Cross-Hemispheric Cross-Validation Tasks | | Leave-One-Trajectory Out Cross-Validation Tasks | | | | |
|---|---|---|---|---|---|---|---|
| | **train**: right **test**: left | **train**: left **test**: right | **train**: non-anterior **test**: anterior | **train**: non-central **test**: central | **train**: non-lateral **test**: lateral | **train**: non-medial **test**: medial | **train**: non-posterior **test**: posterior |
| **balanced accuracy** | 0.633 | 0.610 | 0.633 | 0.583 | 0.649 | 0.673 | 0.574 |
| **weighted-AUC** | 0.699 | 0.690 | 0.713 | 0.655 | 0.698 | 0.763 | 0.661 |
| **weighted-F1** | 0.632 | 0.612 | 0.635 | 0.593 | 0.653 | 0.688 | 0.574 |

**Table 12 – Performances of Trained Classifiers for GPi In-Out Localization in Cross-Validation Tasks**

Instead of evaluating the performance of the selected neural biomarkers in a combined fashion, we also want to understand the effect of individual markers on localization in different spatial domains. Hence, we trained Voting classifiers with single biomarkers individually and check the performances by comparing them with random chance intervals. In this and subsequent sections, we will also share these results.

| Hemisphere | Biomarkers | Balanced Accuracy | | | Weighted AUC | | | Weighted F1 | | |
|---|---|---|---|---|---|---|---|---|---|---|
| | | Mean | Lower Edge of CI | Upper Edge of CI | Mean | Lower Edge of CI | Upper Edge of CI | Mean | Lower Edge of CI | Upper Edge of CI |
| **Merged** | cv | 0.524 | 0.497 | 0.523 | 0.522 | 0.509 | 0.542 | 0.526 | 0.499 | 0.525 |
| | isi_std | 0.515 | 0.492 | 0.520 | 0.530 | 0.503 | 0.538 | 0.517 | 0.494 | 0.522 |
| | delta_band_max_power | 0.522 | 0.473 | 0.503 | 0.538 | 0.477 | 0.513 | 0.524 | 0.476 | 0.505 |
| | delta_band_min_power | 0.576 | 0.487 | 0.514 | 0.589 | 0.493 | 0.527 | 0.579 | 0.489 | 0.516 |
| | theta_band_min_power | 0.550 | 0.486 | 0.513 | 0.571 | 0.492 | 0.525 | 0.552 | 0.488 | 0.515 |
| | alpha_band_min_power | 0.510 | 0.488 | 0.517 | 0.517 | 0.497 | 0.531 | 0.512 | 0.490 | 0.519 |
| | gamma_band_min_power | 0.525 | 0.491 | 0.520 | 0.540 | 0.502 | 0.536 | 0.528 | 0.493 | 0.521 |
| | combination | 0.617 | 0.556 | 0.584 | 0.691 | 0.619 | 0.646 | 0.618 | 0.559 | 0.587 |
| **Right** | firing_rate | 0.488 | 0.489 | 0.528 | 0.475 | 0.501 | 0.548 | 0.489 | 0.490 | 0.530 |
| | cv | 0.562 | 0.494 | 0.540 | 0.597 | 0.514 | 0.566 | 0.563 | 0.496 | 0.541 |
| | isi_mean | 0.508 | 0.503 | 0.539 | 0.490 | 0.520 | 0.565 | 0.509 | 0.504 | 0.542 |
| | delta_band_max_power | 0.509 | 0.471 | 0.508 | 0.548 | 0.473 | 0.520 | 0.508 | 0.472 | 0.509 |
| | theta_band_min_power | 0.604 | 0.489 | 0.529 | 0.630 | 0.497 | 0.547 | 0.607 | 0.491 | 0.530 |
| | beta_band_max_power | 0.552 | 0.483 | 0.523 | 0.552 | 0.492 | 0.538 | 0.552 | 0.484 | 0.525 |
| | delta_band_peak_coherence | 0.602 | 0.487 | 0.521 | 0.631 | 0.508 | 0.547 | 0.599 | 0.455 | 0.495 |
| | delta_band_peak_coherence_frequency | 0.556 | 0.469 | 0.499 | 0.597 | 0.468 | 0.510 | 0.550 | 0.436 | 0.471 |
| | combination | 0.633 | 0.559 | 0.591 | 0.705 | 0.627 | 0.662 | 0.630 | 0.560 | 0.592 |
| **Left** | firing_rate | 0.533 | 0.492 | 0.530 | 0.573 | 0.500 | 0.549 | 0.533 | 0.492 | 0.531 |
| | cv | 0.549 | 0.495 | 0.540 | 0.526 | 0.507 | 0.563 | 0.542 | 0.495 | 0.541 |
| | isi_mean | 0.529 | 0.497 | 0.536 | 0.571 | 0.520 | 0.560 | 0.529 | 0.497 | 0.537 |
| | isi_std | 0.581 | 0.496 | 0.536 | 0.620 | 0.510 | 0.559 | 0.582 | 0.496 | 0.536 |
| | combination | 0.636 | 0.540 | 0.577 | 0.664 | 0.590 | 0.626 | 0.637 | 0.541 | 0.578 |

**Table 13 – Performances of Voting Classifiers Trained with Single and Combined Version of Biomarkers and Random Chance Confidence Intervals for GPi In-Out Localization in Relative Depth Space**

#### 7.2. GPi In-Out Localization in 3D MNI Space

| Biomarker | Merged Hemispheres | | | Right Hemisphere | | | Left Hemisphere | | |
|---|---|---|---|---|---|---|---|---|---|
| | Cramér–Von Mises Test | | Mann-Whitney U Test (corrected) | Cramér–Von Mises Test | | Mann-Whitney U Test (corrected) | Cramér–Von Mises Test | | Mann-Whitney U Test (corrected) |
| | GPi-in | GPi-out | | GPi-in | GPi-out | | GPi-in | GPi-out | |
| **alpha_band_max_power** | x | x | x | x | x | x | 4.340 e-10 | 0.0065 | 0.0011 |
| **alpha_band_mean_coherence** | x | x | x | 0.0349 | 3.674 e-06 | 1.0 | x | x | x |
| **beta_band_mean_coherence** | 6.678 e-10 | 6.246 e-09 | 1.0 | x | x | x | x | x | x |
| **beta_band_mean_significant_coherence** | x | x | x | 0.0010 | 1.875 e-06 | 1.0 | x | x | x |
| **bspike_proportion** | x | x | x | x | x | x | 7.945 e-11 | 1.593 e-10 | 1.0 |
| **cv** | 2.817 e-11 | 4.994 e-10 | 1.0 | x | x | x | x | x | x |
| **delta_band_mean_coherence** | 5.542 e-11 | 4.603 e-11 | 1.0 | 0.0101 | 5.673 e-09 | 1.0 | x | x | x |
| **gamma_band_mean_coherence** | 6.497 e-10 | 4.981 e-07 | 0.8662 | x | x | x | 9.950 e-10 | 0.1726 | 1.0 |
| **gamma_band_peak_coherence** | x | x | x | x | x | x | 0.0484 | 0.1906 | 1.0 |
| **isi_rho** | 0.035 | 0.0096 | 1.0 | x | x | x | 0.0353 | 0.9867 | 0.4913 |
| **isi_skewness** | x | x | x | 0.6793 | 0.0115 | 0.7092 | x | x | x |
| **lv** | 0.0583 | 0.0122 | 1.0 | 0.0143 | 0.0134 | 0.2846 | x | x | x |
| **regularity** | x | x | x | 0.1233 | 0.0052 | 0.7092 | x | x | x |
| **theta_band_mean_coherence** | 2.206 e-11 | 4.187 e-08 | 0.8264 | 0.0092 | 8.436 e-09 | 0.2208 | x | x | x |

**Table 14 – Results of Cramér–Von Mises and Mann-Whitney U Tests Applied on Selected Features for GPi In-Out Localization in 3D MNI Space (p-values are corrected according to Holm-Bonferroni correction)**

| Hemisphere | Performance Metric | Decision Tree Classifier | Random Forest Tree Classifier | K-Nearest Neighbors Classifier | Gaussian Process Classifier | Support Vector Machine Classifier | Voting Classifier (GPC+RF) |
|---|---|---|---|---|---|---|---|
| **merged** | balanced accuracy | 0.543 ±0.041 | 0.596 ±0.038 | 0.577 ±0.022 | 0.557 ±0.031 | 0.556 ±0.043 | 0.600 ±0.033 |
| | weighted-AUC | 0.580 ±0.046 | 0.636 ±0.041 | 0.590 ±0.034 | 0.600 ±0.064 | 0.525 ±0.070 | 0.631 ±0.047 |
| | weighted-F1 | 0.540 ±0.038 | 0.596 ±0.038 | 0.577 ±0.023 | 0.557 ±0.031 | 0.555 ±0.043 | 0.599 ±0.033 |
| **right** | balanced accuracy | 0.616 ±0.113 | 0.673 ±0.130 | 0.646 ±0.060 | 0.695 ±0.048 | 0.650 ±0.097 | 0.705 ±0.117 |
| | weighted-AUC | 0.646 ±0.127 | 0.752 ±0.124 | 0.679 ±0.052 | 0.759 ±0.098 | 0.677 ±0.103 | 0.787 ±0.125 |
| | weighted-F1 | 0.733 ±0.081 | 0.784 ±0.074 | 0.747 ±0.045 | 0.784 ±0.025 | 0.724 ±0.041 | 0.803 ±0.070 |
| **left** | balanced | 0.698 | 0.669 | 0.706 | 0.671 | 0.701 | 0.698 |

| | | | | | | | |
|---|---|---|---|---|---|---|---|
| | accuracy | ±0.087 | ±0.058 | ±0.061 | ±0.086 | ±0.036 | ±0.035 |
| | weighted-AUC | 0.745 ±0.077 | 0.756 ±0.023 | 0.743 ±0.076 | 0.755 ±0.063 | 0.770 ±0.067 | 0.784 ±0.032 |
| | weighted-F1 | 0.782 ±0.046 | 0.743 ±0.024 | 0.742 ±0.061 | 0.785 ±0.044 | 0.754 ±0.052 | 0.790 ±0.028 |

**Table 15 – Performances of Trained Classifiers for GPi In-Out Localization in Main Analysis Modes**

The individual performances of the Decision Tree, Random Forest Tree, KNN, Gaussian Process, and SVM classifiers for GPi in-out localization with the Voting classifier constructed based on soft-voting using the most successful two individual classifiers in MNI space.

| **Performance Metric** | **Cross-Hemispheric Cross-Validation Tasks** | | **Leave-One-Trajectory Out Cross-Validation Tasks** | | | | |
|---|---|---|---|---|---|---|---|
| | **train**: right **test**: left | **train**: left **test**: right | **train**: non-anterior **test**: anterior | **train**: non-central **test**: central | **train**: non-lateral **test**: lateral | **train**: non-medial **test**: medial | **train**: non-posterior **test**: posterior |
| **balanced accuracy** | 0.574 | 0.598 | 0.567 | 0.553 | 0.571 | 0.590 | 0.579 |
| **weighted-AUC** | 0.574 | 0.611 | 0.626 | 0.613 | 0.638 | 0.648 | 0.637 |
| **weighted-F1** | 0.595 | 0.630 | 0.584 | 0.549 | 0.588 | 0.595 | 0.608 |

**Table 16 – Performances of Trained Classifiers for GPi In-Out Localization in Cross-Validation Tasks**

| **Hemisphere** | **Biomarkers** | **Balanced Accuracy** | | | **Weighted AUC** | | | **Weighted F1** | | |
|---|---|---|---|---|---|---|---|---|---|---|
| | | Mean | Lower Edge of CI | Upper Edge of CI | Mean | Lower Edge of CI | Upper Edge of CI | Mean | Lower Edge of CI | Upper Edge of CI |
| **Merged** | isi_rho | 0.512 | 0.467 | 0.512 | 0.545 | 0.517 | 0.554 | 0.530 | 0.478 | 0.530 |
| | cv | 0.531 | 0.488 | 0.534 | 0.547 | 0.502 | 0.552 | 0.521 | 0.510 | 0.541 |
| | lv | 0.534 | 0.498 | 0.543 | 0.549 | 0.493 | 0.551 | 0.529 | 0.492 | 0.534 |
| | beta_band_mean_coherence | 0.519 | 0.482 | 0.539 | 0.535 | 0.501 | 0.537 | 0.511 | 0.482 | 0.519 |
| | theta_band_mean_coherence | 0.489 | 0.465 | 0.518 | 0.508 | 0.485 | 0.529 | 0.496 | 0.485 | 0.531 |
| | delta_band_mean_coherence | 0.517 | 0.476 | 0.527 | 0.540 | 0.487 | 0.546 | 0.495 | 0.491 | 0.517 |
| | gamma_band_mean_coherence | 0.498 | 0.479 | 0.512 | 0.501 | 0.499 | 0.514 | 0.497 | 0.478 | 0.503 |
| | combination | 0.600 | 0.512 | 0.553 | 0.631 | 0.539 | 0.581 | 0.599 | 0.523 | 0.572 |
| **Right** | regularity | 0.530 | 0.491 | 0.543 | 0.626 | 0.499 | 0.574 | 0.736 | 0.572 | 0.607 |
| | lv | 0.521 | 0.501 | 0.553 | 0.626 | 0.510 | 0.588 | 0.728 | 0.580 | 0.613 |
| | isi_skewness | 0.530 | 0.490 | 0.540 | 0.622 | 0.495 | 0.575 | 0.736 | 0.572 | 0.604 |
| | delta_band_mean_coherence | 0.509 | 0.480 | 0.530 | 0.585 | 0.481 | 0.567 | 0.718 | 0.567 | 0.596 |
| | theta_band_mean_coherence | 0.493 | 0.484 | 0.533 | 0.631 | 0.490 | 0.566 | 0.705 | 0.568 | 0.599 |
| | alpha_band_mean_coherence | 0.511 | 0.482 | 0.527 | 0.632 | 0.483 | 0.561 | 0.721 | 0.566 | 0.595 |
| | beta_band_mean_significant_coherence | 0.498 | 0.469 | 0.511 | 0.661 | 0.472 | 0.544 | 0.710 | 0.560 | 0.598 |
| | combination | 0.705 | 0.536 | 0.561 | 0.787 | 0.571 | 0.637 | 0.803 | 0.630 | 0.649 |
| **Left** | isi_rho | 0.500 | 0.474 | 0.526 | 0.592 | 0.495 | 0.568 | 0.731 | 0.645 | 0.667 |
| | bspike_proportion | 0.537 | 0.463 | 0.538 | 0.604 | 0.456 | 0.535 | 0.760 | 0.634 | 0.678 |
| | alpha_band_max_power | 0.502 | 0.474 | 0.526 | 0.696 | 0.453 | 0.561 | 0.733 | 0.635 | 0.667 |
| | gamma_band_mean_coherence | 0.496 | 0.494 | 0.535 | 0.666 | 0.487 | 0.568 | 0.727 | 0.646 | 0.677 |
| | gamma_band_peak_coherence | 0.498 | 0.484 | 0.544 | 0.656 | 0.483 | 0.548 | 0.729 | 0.636 | 0.687 |
| | combination | 0.698 | 0.554 | 0.605 | 0.784 | 0.527 | 0.577 | 0.790 | 0.656 | 0.707 |

**Table 17 – Performances of Voting Classifiers Trained with Single and Combined Version of Biomarkers and Random Chance Confidence Intervals for GPi In-Out Localization in MNI Space**

#### 7.3. GPi Dorsoventral Localization in 3D MNI Space

| Biomarker | Merged Hemispheres | | | Right Hemisphere | | | Left Hemisphere | | |
|---|---|---|---|---|---|---|---|---|---|
| | Cramér–Von Mises Test | | Mann-Whitney U Test | Cramér–Von Mises Test | | Mann-Whitney U Test | Cramér–Von Mises Test | | Mann-Whitney U Test |
| | GPi-in | GPi-out | | GPi-in | GPi-out | | GPi-in | GPi-out | |
| **cv** | 9.338 e-06 | 1.232 e-07 | 0.3391 | 01004 | 0.0163 | 0.0192 | 4.221 e-05 | 2.342 e-06 | 0.8422 |
| **delta_band_min_power** | x | x | x | 0.0082 | 0.0034 | 0.5722 | x | x | x |
| **delta_band_peak_coherence** | 0.0018 | 9.089 e-09 | 0.1883 | x | x | x | x | x | x |
| **firing_rate** | 1.129 e-10 | 3.033 e-11 | 0.0348 | 3769 e-05 | 0.0007 | 0.1219 | 5.358 e-06 | 3.960 e-08 | 0.0825 |
| **isi_mean** | 0.0136 | 1.341 e-05 | 0.0348 | 0.4152 | 0.0179 | 0.1219 | x | x | x |
| **isi_std** | 0.0163 | 1.341 e-05 | 0.0118 | 0.3043 | 0.0499 | 0.0295 | 0.0248 | 5.762 e-05 | 0.0825 |

**Table 18 – Results of Cramér–Von Mises and Mann-Whitney U Tests Applied on Selected Features for GPi Dorsoventral Localization in 3D MNI Space (p-values are corrected according to Holm-Bonferroni correction)**

| Hemisphere | Performance Metric | Decision Tree Classifier | Random Forest Tree Classifier | K-Nearest Neighbors Classifier | Gaussian Process Classifier | Support Vector Machine Classifier | Voting Classifier (KNN+RF) |
|---|---|---|---|---|---|---|---|
| **merged** | balanced accuracy | 0.506 ±0.030 | 0.546 ±0.055 | 0.628 ±0.056 | 0.559 ±0.045 | 0.568 ±0.052 | 0.649 ±0.042 |
| | weighted-AUC | 0.517 ±0.058 | 0.602 ±0.054 | 0.690 ±0.059 | 0.603 ±0.035 | 0.532 ±0.101 | 0.676 ±0.057 |
| | weighted-F1 | 0.538 ±0.028 | 0.563 ±0.049 | 0.638 ±0.054 | 0.586 ±0.039 | 0.585 ±0.049 | 0.661 ±0.039 |
| **right** | balanced accuracy | 0.603 ±0.084 | 0.689 ±0.069 | 0.648 ±0.102 | 0.574 ±0.075 | 0.580 ±0.088 | 0.722 ±0.038 |
| | weighted-AUC | 0.708 ±0.031 | 0.728 ±0.096 | 0.745 ±0.094 | 0.587 ±0.084 | 0.511 ±0.167 | 0.765 ±0.062 |
| | weighted-F1 | 0.562 ±0.086 | 0.685 ±0.072 | 0.637 ±0.108 | 0.548 ±0.088 | 0.575 ±0.075 | 0.712 ±0.047 |
| **left** | balanced accuracy | 0.508 ±0.018 | 0.564 ±0.063 | 0.673 ±0.073 | 0.587 ±0.100 | 0.582 ±0.082 | 0.684 ±0.056 |
| | weighted-AUC | 0.550 ±0.053 | 0.615 ±0.076 | 0.723 ±0.065 | 0.621 ±0.129 | 0.517 ±0.119 | 0.717 ±0.090 |
| | weighted-F1 | 0.559 ±0.015 | 0.608 ±0.055 | 0.690 ±0.062 | 0.623 ±0.087 | 0.620 ±0.064 | 0.704 ±0.053 |

**Table 19 – Performances of Trained Classifiers for Dorsoventral Localization in Main Analysis Modes**

The individual performances of the Decision Tree, Random Forest Tree, KNN, Gaussian Process, and SVM classifiers for dorsoventral localization along with the Voting classifier that constructed based on soft-voting using the most successful two individual classifiers in MNI space.

| Performance Metric | Cross-Hemispheric Cross-Validation Tasks | | Leave-One-Trajectory Out Cross-Validation Tasks | | | | |
|---|---|---|---|---|---|---|---|
| | **train**: right **test**: left | **train**: left **test**: right | **train**: non-anterior **test**: anterior | **train**: non-central **test**: central | **train**: non-lateral **test**: lateral | **train**: non-medial **test**: medial | **train**: non-posterior **test**: posterior |
| **balanced accuracy** | 0.593 | 0.627 | 0.545 | 0.588 | 0.629 | 0.500 | 0.601 |
| **weighted-AUC** | 0.651 | 0.718 | 0.578 | 0.677 | 0.722 | 0.653 | 0.711 |
| **weighted-F1** | 0.628 | 0.629 | 0.554 | 0.672 | 0.654 | 0.499 | 0.614 |

**Table 20 – Performances of Trained Classifiers for Dorsoventral Localization in Cross-Validation Tasks**

| Hemisphere | Biomarkers | Balanced Accuracy | | | Weighted AUC | | | Weighted F1 | | |
|---|---|---|---|---|---|---|---|---|---|---|
| | | Mean | Lower Edge of CI | Upper Edge of CI | Mean | Lower Edge of CI | Upper Edge of CI | Mean | Lower Edge of CI | Upper Edge of CI |
| **Merged** | cv | 0.602 | 0.494 | 0.530 | 0.633 | 0.510 | 0.562 | 0.636 | 0.513 | 0.548 |
| | delta_band_peak_coherence | 0.554 | 0.486 | 0.511 | 0.579 | 0.504 | 0.547 | 0.593 | 0.491 | 0.521 |
| | firing_rate | 0.594 | 0.481 | 0.517 | 0.623 | 0.490 | 0.543 | 0.626 | 0.499 | 0.534 |
| | isi_mean | 0.552 | 0.490 | 0.526 | 0.565 | 0.513 | 0.559 | 0.594 | 0.508 | 0.544 |
| | isi_std | 0.618 | 0.491 | 0.523 | 0.658 | 0.503 | 0.550 | 0.660 | 0.510 | 0.541 |
| | combination | 0.649 | 0.543 | 0.565 | 0.676 | 0.599 | 0.644 | 0.661 | 0.559 | 0.583 |
| **Right** | isi_std | 0.572 | 0.549 | 0.579 | 0.649 | 0.557 | 0.593 | 0.555 | 0.543 | 0.573 |
| | cv | 0.595 | 0.534 | 0.563 | 0.659 | 0.546 | 0.582 | 0.590 | 0.528 | 0.556 |
| | firing_rate | 0.711 | 0.519 | 0.547 | 0.760 | 0.535 | 0.568 | 0.713 | 0.512 | 0.539 |
| | isi_mean | 0.757 | 0.550 | 0.577 | 0.764 | 0.554 | 0.587 | 0.756 | 0.543 | 0.571 |
| | delta_band_min_power | 0.601 | 0.528 | 0.560 | 0.562 | 0.536 | 0.576 | 0.589 | 0.519 | 0.553 |
| | combination | 0.722 | 0.555 | 0.583 | 0.765 | 0.585 | 0.618 | 0.712 | 0.549 | 0.577 |
| **Left** | isi_std | 0.596 | 0.538 | 0.553 | 0.618 | 0.554 | 0.571 | 0.651 | 0.592 | 0.606 |
| | cv | 0.559 | 0.534 | 0.552 | 0.586 | 0.552 | 0.570 | 0.611 | 0.594 | 0.606 |
| | firing_rate | 0.569 | 0.521 | 0.537 | 0.528 | 0.532 | 0.553 | 0.602 | 0.577 | 0.592 |
| | combination | 0.684 | 0.580 | 0.592 | 0.717 | 0.604 | 0.622 | 0.704 | 0.639 | 0.650 |

**Table 21 – Performances of Voting Classifiers Trained with Single and Combined Version of Biomarkers and Random Chance Confidence Intervals for GPi Dorsoventral Localization in MNI Space**

| Patient ID | BFMDRS | | | | |
|---|---|---|---|---|---|
| | Arm | | Leg | | |
| | Baseline Right Arm | Baseline Left Arm | Baseline Right Leg | Baseline Left Leg | More Affected Side |
| **1** | 2 | 2 | 0 | 0 | **Equal** |
| **2** | 6 | 0 | 6 | 0 | **Right** |
| **3** | 6 | 6 | 12 | 12 | **Equal** |
| **4** | 9 | 9 | 16 | 0 | **Right** |
| **5** | x | x | x | x | **x** |
| **6** | 0 | 0 | 0 | 0 | **Equal** |
| **7** | x | x | x | x | **x** |
| **8** | 0 | 0 | 12 | 4 | **Right** |
| **9** | 4 | 4 | 12 | 0 | **Right** |
| **10** | 1 | 0 | 4 | 0 | **Right** |

**Table 22 – The Baseline BFMDRS Scores of Arms and Legs.**
